# Impact of oseltamivir treatment on influenza A and B dynamics in human volunteers

**DOI:** 10.1101/2020.11.13.20231357

**Authors:** Kyla L. Hooker, Vitaly V. Ganusov

## Abstract

Influenza viruses infect millions of humans every year causing an estimated 400,000 deaths globally. Due to continuous virus evolution current vaccines provide only limited protection against the flu. Several antiviral drugs are available to treat influenza infection, and one of the most most commonly used drugs is oseltamivir (Tamiflu). While the mechanism of action of oseltamivir as a neuraminidase inhibitor is well understood, the impact of oseltamivir on influenza virus dynamics in humans has been controversial. Many clinical trials with oseltamivir have been done by pharmaceutical companies such as Roche but the results of these trials until recently have been reported as summary reports or papers. Typically, such reports included median virus shedding curves for placebo and drug-treated influenza virus infected volunteers often indicating high efficacy of the early treatment. However, median shedding curves may be not accurately representing drug impact in individual volunteers. Importantly, due to public pressure clinical trials data testing oseltamivir efficacy has been recently released in the form of redacted PDF documents. We digitized and re-analyzed experimental data on influenza virus shedding in human volunteers from three previously published trials: on influenza A (1 trial) or B viruses (2 trials). Given that not all volunteers exposed to influenza viruses actually start virus shedding we found that impact of oseltamivir on the virus shedding dynamics was dependent on i) selection of volunteers that were infected with the virus, and ii) the detection limit in the measurement assay; both of these details were not well articulated in the published studies. By assuming that any viral measurement is above the limit of detection we could match previously published data on median influenza A virus (flu A study) shedding but not on influenza B virus shedding (flu B study B) in human volunteers. Additional analyses confirmed that oseltamivir had an impact on the duration of shedding and overall shedding (defined as area under the curve) but this result was varied by the trial. Interestingly, treatment had no impact on the rates at which shedding increased or declined with time in individual volunteers. Additional analyses showed that oseltamivir impacted the kinetics of the start and end of viral shedding and in about 20-40% of volunteers treatment had no impact on viral shedding duration. Our results suggest an unusual impact of oseltamivir on influenza viruses shedding kinetics and caution about the use of published median data or data from a few individuals for inferences. Furthermore, we call for the need to publish raw data from critical clinical trials that can be then independently analyzed.

## Introduction

Influenza is a respiratory infection caused by different strains of the influenza virus. Influenza A viruses originate from animals such as birds and pigs while influenza B viruses have no known animal origin [1]. Disease, caused by influenza viruses, commonly known as the flu, typically affects the upper respiratory system such as the sinus cavities, throat, and sometimes the lungs [2]. The virus spreads from person to person via respiratory droplets when the infected individual coughs or sneezes in close contact with uninfected individuals [3]. Symptoms include fever, fatigue, cough, sore throat, and a runny nose. Most individuals recover from the flu [2]. However, individuals, usually with underlying health conditions, can have serious and even deadly complications. Millions are infected with influenza viruses gobally, and 400,00-500,000 people die each year from complications following influenza virus infections [1, 4]. The influenza virus has a high mutation rate resulting in new strains (antigenic drift) that are not readily recognized by immunity of individuals previously experienced influenza infection [2]. Occasionally, reassortment of viral genes may occur resulting in variants that are markedly different from currently circulating strains (antigenic shift); such process often results in a pandemic [5].

A common prevention is the yearly flu vaccine, but it is not very efficient with an estimated efficacy of about 60% which varies with vaccination year and age of vaccinated individuals [6]. The influenza virus has a high mutation rate allowing it to escape from vaccine-induced immunity [2]. Thus, new influenza vaccines need to be created annually. The creation of the annual influenza vaccination takes into consideration both new strains and current strains of influenza viruses that are circulating globally [1, 7].

Luckily there are several antiviral drugs such as oseltamivir, zanamivir, peramivir, and baloxavir that can be used to either treat severely ill influenza-infected patients or household contacts of individuals with confirmed influenza infection [8–10]. Efficacy of such drugs has been extensively evaluated in clinical trials both including infected patients and volunteers that had been infected with a known strain and dose of an influenza virus. In latter types of experiments treatment start can be well defined relative to the infection initiation, and for oseltamivir the treatment appears to impact virus dynamics and/or patient’s symptoms when the treatment is started only within 24 hours of the infection and/or onset of symptoms [2]. There are also other limitations of oseltamivir including side effects and the appearance of drug resistant variants [2, 11].

While evidence of the efficacy of drugs against influenza infection such as oseltamivir has been well documented from several clinical trials (e.g., [12, 13]) for a long time there has been very limited publicly available data from such clinical trails. In particular, results of clinical trials have been presented as median viral shedding curves or symptoms for placebo and drug-treated volunteers, and side effects of the treatments were barely discussed. Interestingly, initial reviews of such clinical trials data recommended oseltamivir use for treating influenza infection [14]; many governments stockpiled oseltamivir for emergency use in case of a new pandemic virus [15]. However, concerns of whether the clinical trials data were accurately represented in original publications were raised resulted in some reports of the clinical trials with oseltamivir to be publicly released. Interestingly, reanalysis of these and other data reduced the initial enthusiasm to recommend oseltamivir for routine treatment of uncomplicated influenza infections [16, 17]. While the data from several of the early clinical trials are now available, these data are given as pdf scans of redacted reports and not the actual raw data which precludes more detailed analysis by other investigators. Moreover, as far as we are aware data from most recent clinical trials of other anti-influenza drugs such as baloxavir are not publicly available [18].

Having properly formatted, digitized data from clinical trials could be extremely useful to understanding the impact of the drug treatment on influenza virus dynamics in humans. Furthermore, such shedding data could be useful to further understand mechanisms that control duration and magnitude of viral shedding in humans. For example, kinetics of influenza A virus shedding in several human volunteers has been analyzed with the use of mathematical models [19–21]. Interestingly, one of the earliest modelling studies suggested that the dynamics of influenza A virus shedding in human volunteers can be well described by a so-called target cell limited model in which virus dynamics is only restricted by the availability of targets for virus replication [19]. However, Baccam *et al*. [19] study used data from only a few volunteers that were not treated with drugs; therefore, it remains unclear if the same model can describe more variable data from a large group of infected volunteers, or if other alternative models of viral control may be also consistent with viral shedding patterns [22, 23].

In this paper we carefully digitized data on influenza virus shedding in human volunteers from three previously published clinical trials and performed basic analysis of these data. The primary goals of the analysis were to reproduce published results on oseltamivir treatment impact on viral shedding and to provide the community with well curated datasets on viral dynamics that other researchers may utilize further.

## Material and methods

### Experimental Design

Three randomized placebo-controlled clinical trials using influenza A virus (H1N1, 1 trial) or influenza B virus (2 trials) were done using volunteers [12, 13]. In short, volunteers were inoculated through the nose with the influenza virus strain of the trial at time zero. Treatment type varies by trial (Figure 1 and Table 1). The influenza A trial used a placebo group and a group treated with oseltamivir at daily with 20 mg, 100 mg, 200 mg bid (twice daily), or 200 mg od (once daily). Influenza B study A trial used a placebo group and a group treated daily with oseltamivir at 75 mg or 150 mg. Influenza B study B trial used a placebo group and a group treated daily with 75 mg of oseltamivir. In all three trials treatment began at 24 hours after virus inoculation and nasal washings were taken every 12 hours until 96 hours (4 days) after inoculation. At that point nasal washings were taken every 24 hours until 216 hours (9 days). The nasal washings were tested for the viral titer (amount of viral shedding in the patient).

**Table 1:** Details of the datasets analyzed in the paper. Some volunteers were excluded from the analysis because the volunteer’s viral titers never exceeded the limit of detection, and thus were interpreted as uninfected. Impact of the limit of detection (LOD) on the number of patients selected as “infected” for every study is evaluated in Table 2. Data are from previously published clinical trials [12, 13]. Other used abbreviations: bid: twice daily, od: once daily.

| Study | LOD<br>TCID/ml | # of all<br>volunteers | # of placebo volunteers:<br>Included[Excluded] | # of treated volunteers: Included[Excluded] |  |  |  |  |  |
| --- | --- | --- | --- | --- | --- | --- | --- | --- | --- |
|  |  |  |  | 20mg | 75mg bid | 100mg | 150mg bid | 200mg bid | 200mg od |
| FluA | 0 | 80 | 12[4] | 13[3] | - | 13[3] | - | 15[1] | 13[3] |
| FluB study A | 0 | 60 | 13[7] | - | 10[10] | - | 10[10] | - | - |
| FluB study B | 0 | 117 | 27[12] | - | 49[29] | - | - | - | - |

**Figure 1:**
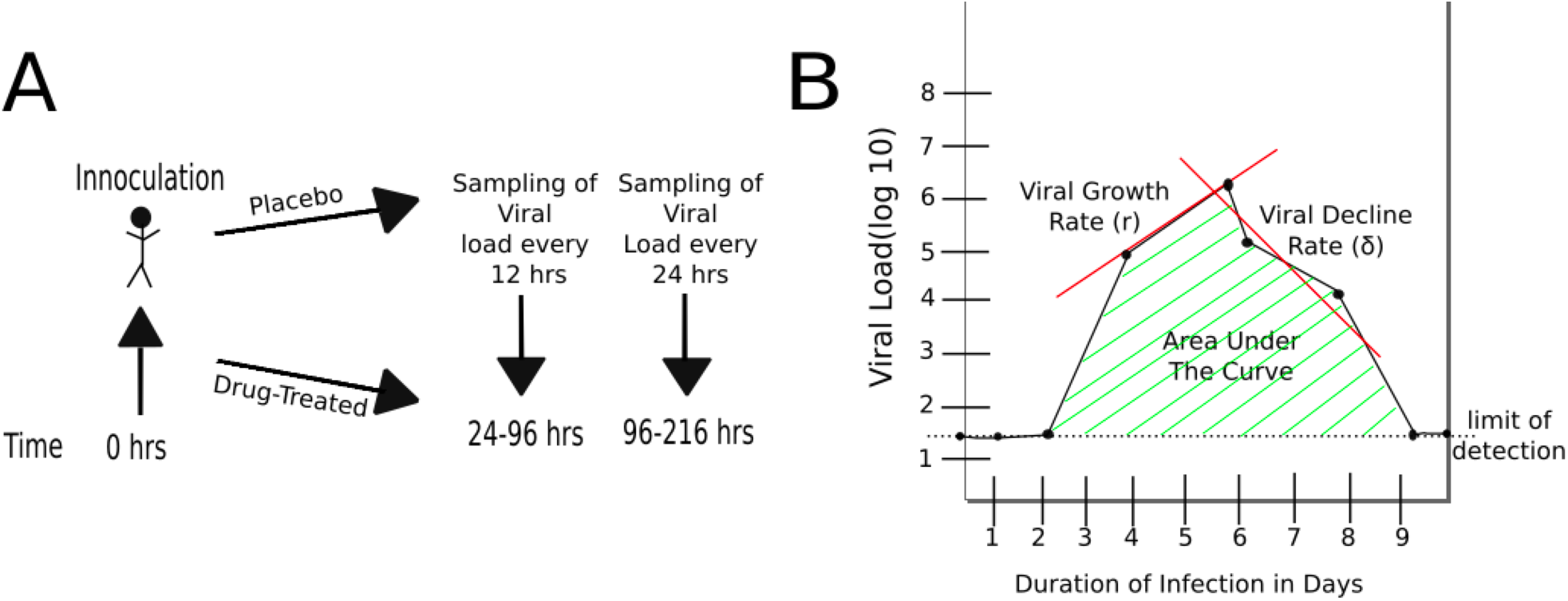
Schematic representation of experimental design and basic characteristics of viral shedding data. A) volunteers were inoculated with influenza A or B virus at time zero, and treatment with oseltamivir (or placebo) started at 24 hours [12, 13]. Nasal washing to measure viral titers were taken every 12 hours starting at 24 hours until 96 hours, and every 24 hours thereafter until 216 hours (9 days). B) Basic parameters estimated from kinetics of shedding including the duration of infection (DOI or *T*; *T* = 8 days in the cartoon), total viral shedding (area under the curve, AUC), viral growth rate (*r*), and viral decline rate (*δ*). Area under the curve (AUC) was calculated for viral concentration on the linear scale and then log_10_-transformed (see eqn. (1)).

### Experimental Data

#### Data digitization

Redacted PDF files describing in detail clinical trials have been downloaded in 2014 from dryad.org (https://datadryad.org/resource/doi:10.5061/dryad.77471/2). Data from three trials involving treatment of volunteers with oseltamivir (Tamiflu) 24 hours after controlled exposure to influenza viruses were chosen for further analysis: study on infection with influenza A virus (“Flu A study”, report `PV15616`), and two studies on infection with influenza B virus (“Flu B study A”, report `NP15717` and “Flu B study B”, report `NP15827`) [12, 13]. The data in the original PDF files were given in day:hour:min time units for times of virus inoculation or when measurements of viral shedding were taken. Viral titers were measured in tissue-culture infectious doses per ml (TCID_50_*/*ml) and are given in log_10_ units. The data were digitized by KLH into a spreadsheet format with time given in minutes or days since infection. Accuracy of digitization was confirmed by checking the correspondence between pdf files by another student for a set of randomly chosen volunteers.

#### Defining uninfected volunteers

Not all virus-exposed volunteers shed the virus after virus inoculation. Volunteers with no viral titer at any point during the trial were excluded from the data analysis (see (Table 1 and Figures S1-S8 for the number and the lists of excluded volunteers).

#### Calculating start and stop of shedding

To calculate the kinetics at which volunteers started virus shedding in a given cohort (e.g., placebo-treated individuals in Flu A study) we did the following. First, for each volunteer we converted the shedding data into 0 or 1 with 0 values being assigned for times when viral shedding was at or below the LOD and 1 values assigned for all times when viral shedding exceeded LOD or the time was later than the first time point of the positive viral shedding event. Then we used equally spaced time points (0, 0.5, 1, 1.5, etc. days) and we counted the number of 0 or 1 for all volunteers in the cohort. The resulting data are given as the number of volunteers that started shedding the virus at time *t* = 0, 0.5, 1 … days after the virus inoculation. Similarly, to calculate the time by which volunteers stop shedding we similarly converted viral shedding data to 0 or 1 but starting counting time in reverse, starting with the latest time point going backwards. We then similarly calculated the number of individuals in the cohort that are still shedding the virus by time *t* where *t* is from sequence 0, 0.5, 1, 1.5, etc. days point infection. The resulting data are given as the number of volunteers that were still shedding the virus at time *t* = 0, 0.5, 1 … days after the virus inoculation. Generated data for each of the clinical trials are provided as supplement.

#### Median data from published studies

To compare median shedding curves calculated in our datasets with published values we digitized data for influenza A virus shedding (from Figure 3 in Hayden *et al*. [12]) or influenza B virus shedding (Flu B study B from Figure 2 in Hayden *et al*. [13]). Data were digitized using Engauge Digitizer and are available as supplement to this paper.

**Figure 2:**
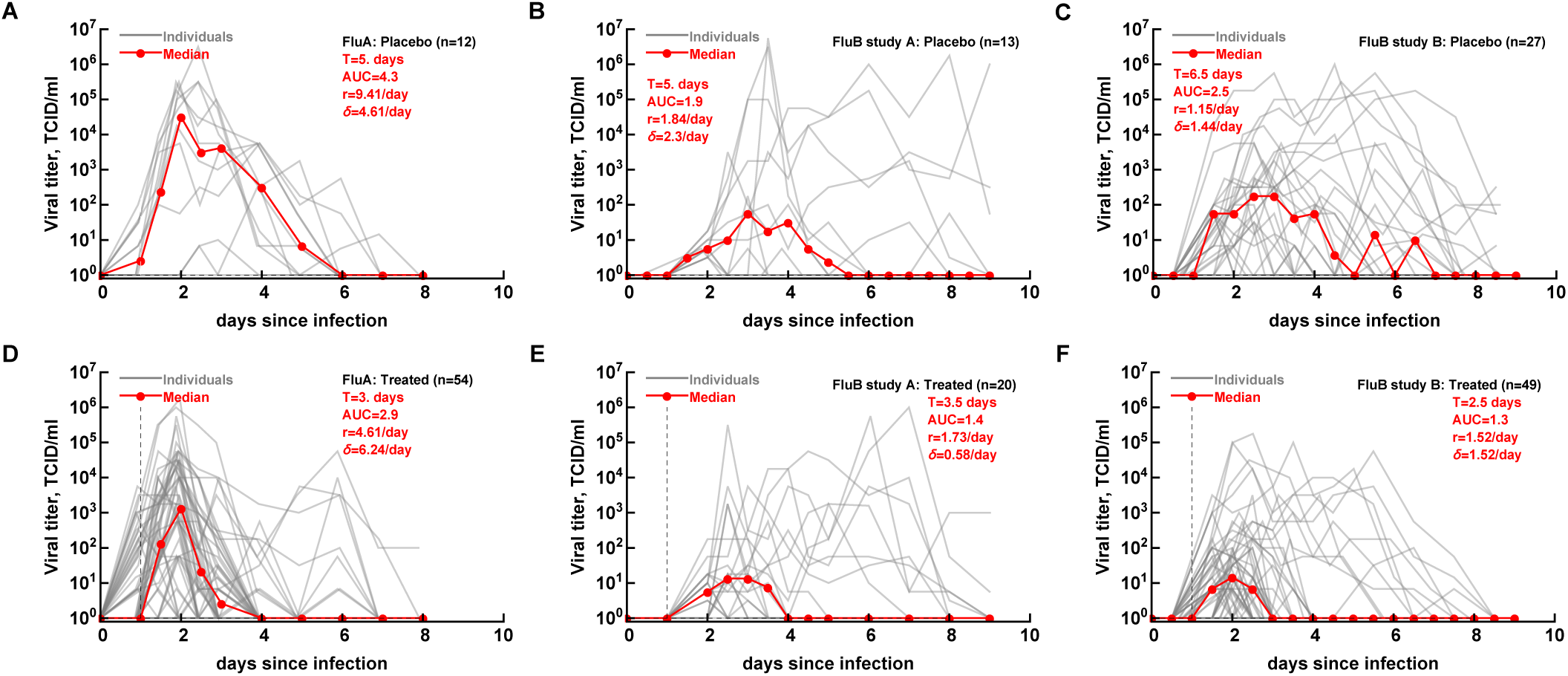
Commonly reported median viral shedding curves do not accurately represent shedding in many patients. We analyzed experimental data from 3 clinical trials of volunteers infected with influenza A (panels A&D) or influenza B (panels B-C&E-F) viruses and treated 24 hours after viral exposure with placebo (panels A-C) or oseltamivir (panels D-F); start of treatment is indicated by the vertical dashed lines. Viral shedding in individual volunteers is shown by gray lines and median viral titers are shown by thick red lines with markers. For median viral titers we also calculated the duration of infection (*T*), the rate of viral growth and viral decay (*δ*), and the total area under the curve (AUC, eqn. (1)). Details of the experiments and basic viral characteristics calculated are given in [12, 13, see Figure 1]. The limit of detection is represented by the horizontal thin dashed line; the detection limit was 0 log_10_ TCID_50_*/*ml for all data. Viral shedding curves in individual volunteers are shown in Supplement (Figures S1–S9).

**Figure 3:**
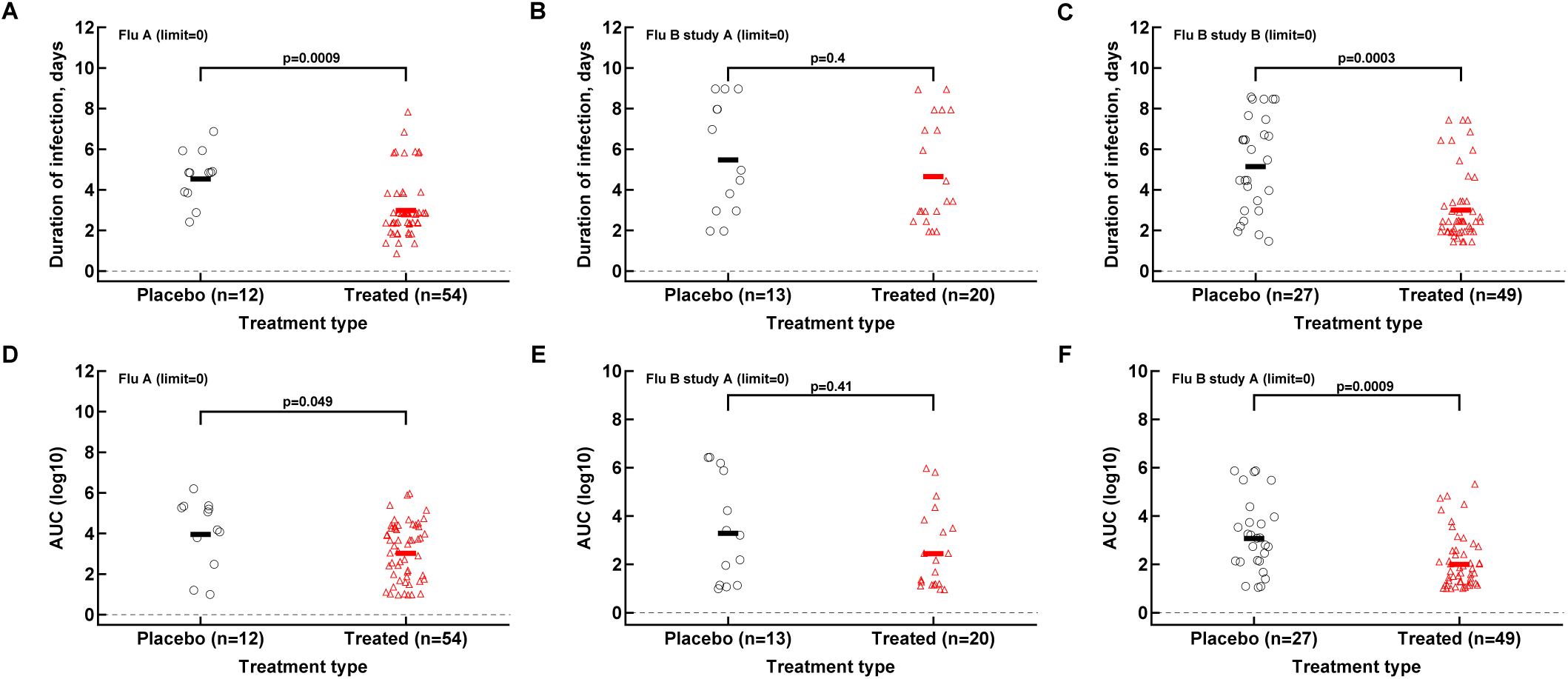
Impact of oseltamivir treatment on the duration of infection and the overall viral shedding depends on the study. For every volunteer in 3 clinical trials we calculated the duration of infection (panels A-C) or the overall shedding defined as AUC (see Materials and methods for detail) for Flu A (A&D), Flu B study A (B&E), or Flu B study B (C&F) data. Horizontal lines indicate median values. The number of volunteers, *n*, analyzed in each of the trial and tests is shown on the *x*-axis of each graph. Comparisons between groups done using Mann-Whitney test and *p* values from the test are shown on individual panels.

### Statistical Analysis

#### Duration of infection (DOI)

Duration of infection was defined as the last time point at which shedding was above the limit of detection.

#### Area under the curve (AUC)

To calculate AUC we converted viral titers which were given in log_10_ units to the linear scale and then calculated AUC using trapezoid integration method:

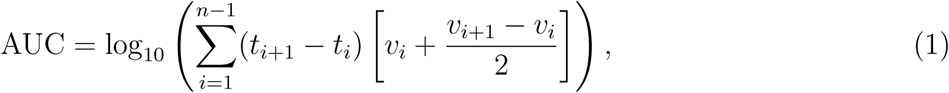

where *t*_*i*_ is the *i*^*th*^ time point of measurement of viral load, 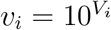 where *V*_*i*_ is the viral load in the data, and *n* is the total number of time points.

#### Viral shedding increase (*r*) or decline (*δ*) rates

To calculate the exponential increase (*r*) in the viral shedding we selected shedding data which are above the LOD and up to the peak of shedding. The growth rate was calculated as the slope of linear function fitted to these (log10) viral titer data multiplied by ln(10) (*r* = slope × ln(10)). In a similar fashion, for calculating the exponential decline rate (*δ*) in the viral shedding we selected data after the peak of viral shedding (including the peak value) until the last shedding value above the LOD. The early growth rate was calculated using the last measurement at the LOD and all measurements above the LOD including before the maximum viral titer measurement. In a similar fashion, the late decline rates included all points after the maximum viral titer that were above the LOD and the first measurement at the LOD.

#### Mathematical modeling of viral shedding start/end

To quantify the rates at which volunteers start or stop shedding we used a novel mathematical model. In the model we assume that the population of shedders may consist of two sub-populations with fraction *f* and 1 *- f* and each population either start or stop shedding at rates *s*_1_ and *s*_2_ respectively. We assume that progression of a volunteer from “non-shedding” to “shedding” state occurs as a movement via *k* subcompartments at a rate *s*_1_ or *s*_2_ [24]. Then the probability that a volunteer starts shedding the virus at time *t* is given by an incomplete gamma function 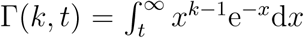 where *k* is the shape parameter of the distribution and Γ(*k*) = Γ(*k*, 0) = (*k*−1)!. Assuming that shedding starts (or stops) after a delay *τ* the proportion of volunteers that start shedding by time *t* after infection is given by the formula

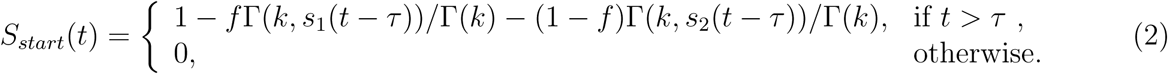

To describe how volunteers stop shedding the virus at time *t* after infection we used formula

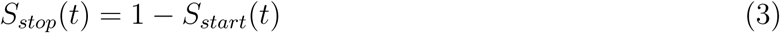

with *S*_*start*_(*t*) being defined in eqn. (2). To characterize the speed at which volunteers start or stop shedding we used average time defined as *T*_*i*_ = *k/s*_*i*_. To fit the models to experimental data we used a likelihood approach that had been used previously to describe viral escape from T cell immunity [25, 26]. Specifically, to describe the fraction of volunteers that start shedding the virus the negative log-likelihood of the data given the model is

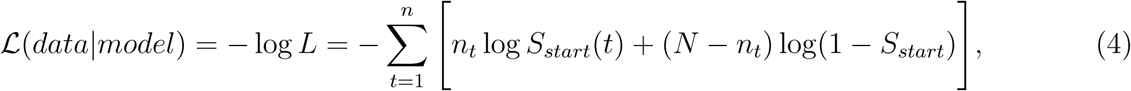

where *n*_*t*_ is the number of volunteers who have started virus shedding by time *t, N* is the total number of volunteers in the cohort, *S*_*start*_ is given in eqn. (2), and we ignored the constant terms which are irrelevant when maximizing likelihood. Alternative models in this analysis also included one population model (*f* = 1 and *s*_2_ = 0), no delay (*τ* = 0), or exponentially distributed shedding times (*k* = 1). Confidence intervals for the model parameters were estimated using bootstrap approach [27]. For each cohort we resampled volunteers in the cohort with replacement and then followed the same procedure outlined above and calculated the number of individuals that start (or stop) shedding by particular time after the virus inoculation. The model was then fit to 1,000 of such resampled datasets for each cohort, and 95% confidence intervals were calculated from the distributions of estimated parameters.

#### Tests

All major analyses were done in R (version 3.1) or Mathematica 11.3. Fitting the mathematical models to data was done in Mathematica 11.3. Statistical comparisons for various parameters estimated for placebo and treated volunteers were done using nonparameteric unpaired Wilcoxon test (identical to Mann-Whitney test). Nested models were compared using likelihood ratio test (LRT). To compare similarity in kinetics of start or end of virus shedding between placebo- and drug-treated volunteers we fitted the data from two cohorts with either individual parameters per cohort/dataset or with the same parameters for both cohorts/datasets. We then used likelihood ratio test to determine if parameters for virus shedding start/end were different between the two cohorts/datasets.

## Results

### Defining limit of detection

In clinical trials testing efficacy of oseltamivir, healthy volunteers were inoculated with a defined dose of the influenza A or B viruses (see Materials and methods for more detail and [12, 13]). Interestingly, many volunteers did not shed any detectable virus for the whole duration of the clinical trial (17.5% in flu A clinical trial, 45% in flu B study A and 35% in flu B study B, Figures S1–S9). These individuals were excluded from further analyses on the kinetics of viral shedding. While most other volunteers showed consistent and high viral shedding, some volunteers showed detectable (above zero) shedding only at one time point. It was unclear from the study descriptions whether such individuals should be counted as infected or if such viral blips are false positives. We investigated how changing the limit of detection (LOD) may impact the number of volunteers classified as infected for these clinical trials.

We reasoned that because treatment started 24 hours after infection and replication cycle of influenza viruses is relatively short (< 24 h [28, 29]), oseltamivir should not impact the probability of a person to be infected [30]; therefore, by changing the LOD the difference between percent of infected in placebo- or drug-treated groups can be evaluated using two-by-two contingency tables (Table 2). The basic idea was to vary the LOD to several values that viral titers take in a given trial and see at which values the frequency of volunteers that shed the virus at any time point (i.e., have viral titers above the threshold value) is the same between placebo and treatment groups. We tried several different values of the LOD and only in 3rd trial (Flu B study B) increasing the LOD to 1.5 log_10_ TCID_50_*/*ml impacted significantly the frequency of infection between placebo and treated volunteers. Because increasing LOD reduced the number of volunteers shedding the virus above LOD (Table 2), we converged to use LOD = 0 in our further calculations; some results were also checked assuming other values for LOD (see below). Importantly, using LOD = 0 we could match the median virus shedding titers for placebo- and drug-treated volunteers in Flu A study suggesting that this was the likely LOD in that study (see below).

**Table 2:** Impact of the limit of detection (LOD) on the number of volunteers defined as infected or uninfected in three analyzed clinical trials. We used different values for LOD to define which individuals became infected following exposure to influenza viruses. Individual was defined infected when viral shedding was above LOD at least at one time point during the trial. The number of included/excluded individuals is shown in individual columns. We also performed a 2×2 contingency table test by comparing the fraction of infected individuals in placebo vs. treated groups, and the the p-values from the tests (both 1 tailed and 2 tailed) are shown.

| Influenza A<br>LOD | # Placebo Excluded | # Placebo Included | # Treated Excluded | # Treated Included | P-value<br>(1-tailed/2-tailed) |
| --- | --- | --- | --- | --- | --- |
| 0.0 | 0 | 12 | 0 | 54 | 1.0/1.0 |
| 0.84 | 1 | 11 | 6 | 48 | 0.63/1.0 |
| 1.76 | 2 | 10 | 13 | 41 | 0.45/0.72 |
| 2.76 | 4 | 8 | 24 | 30 | 0.36/0.54 |
| Influenza B Study A<br>LOD | # Placebo Excluded | # Placebo Included | # Treated Excluded | # Treated Included | P-value<br>(1-tailed/2-tailed) |
| 0.0 | 0 | 13 | 0 | 21 | 1.0/1.0 |
| 0.50 | 1 | 12 | 3 | 18 | 0.50/1.0 |
| 1.0 | 4 | 9 | 6 | 15 | 0.59/1.0 |
| Influenza B Study B<br>LOD | # Placebo Excluded | # Placebo Included | # Treated Excluded | # Treated Included | P-value<br>(1-tailed/2-tailed) |
| 0.0 | 0 | 27 | 0 | 49 | 1.0/1.0 |
| 0.42 | 0 | 27 | 0 | 27 | 1.0/1.0 |
| 0.84 | 3 | 24 | 9 | 40 | 0.32/0.52 |
| 1.50 | 4 | 23 | 21 | 28 | 0.011/0.021 |

### Impact of oseltamivir treatment on overall shedding pattern in human volunteers

Having defined the LOD for the data we then compared how accurately median shedding curves represent virus shedding patterns in individual volunteers (Figure 2; see Materials and Methods for how median shedding curves were calculated). Interestingly, for flu A-infected volunteers there was a reasonable correspondance between median shedding curves and shedding for individual placebo- and drug-treated volunteers; however, there are clearly examples of individuals that did not follow the median pattern (Figure 2A&D). The match between median shedding and shedding in individual volunteers was poor for both studies with influenza B virus; particularly, median curves poorly represented individuals that continued to shed the virus in both placebo- and drug-treated controls (e.g., Figure 2B&E) illustrating a severe limitation of presenting the data by median shedding alone.

For our median shedding curves we calculated several basic parameters characterizing virus dynamics such as the duration of infection (*T*), the overall viral shedding (defined as area under the curve, AUC), the virus growth rate prior to peak *r*, and the virus decline rate after the peak *δ*. Treatment impacted these parameters for the median shedding curves differently. In particular, oseltamivir treatment reduced the median duration of infection from 5 to 3 days and reduced the overall viral shedding over 10 fold (from 4.2 to 2.9) in flu A clinical trial as judged by the median shedding curves (Figure 2A&D). Also as judged by the median shedding curves, in influenza A virus-infected individuals treatment reduced the rate of viral growth and increased the rate of viral clearance after the peak (Figure 2A&D). In contrast, in flu B study A drug treatment did not reduce the overall median shedding and resulted in slower viral clearance after the peak (Figure 2B&E). Finally, in flu B study B, treatment did not appear to have an impact on viral growth or decline rates of the median shedding data but did result in shorter infection and less overall virus being shed (Figure 2C&F).

Given that median viral shedding curves did not necessarily well represent shedding observed in individual volunteers, impact of the treatment on median shedding curves may be misleading. Therefore, we performed an alternative analysis in which we calculated the same parameters (*T*, AUC, *r*, and *δ*) for individual patients (Figure 3). It is important to emphasize that it was not possible to calculate virus growth and decay rates for all volunteers because this procedure required at least two measurements to be above LOD during viral increase or decline phases which was not available in all volunteers. Interestingly, we found that oseltamivir treatment did reduce the overall duration of infection and the total shedding in flu A study and flu B study B, but there was no difference in these two parameters between placebo- and drug-treated individuals in flu B study A (Figure 3).

Our analysis of the virus kinetics defined by the virus shedding growth rate to the peak and virus decline rate after the peak based on median virus shedding curves suggested that oseltamivir influences these rates albeit in trial-dependent manner [22, Figure 2]. Specifically, we found that oseltamivir increases the rate of virus decline after the peak in the median data in flu A clinical trial [22]. In contrast, analysis of individual shedding curves did not reveal impact of the olsetamivir on either viral growth or viral decline rates (Figure 4).

**Figure 4:**
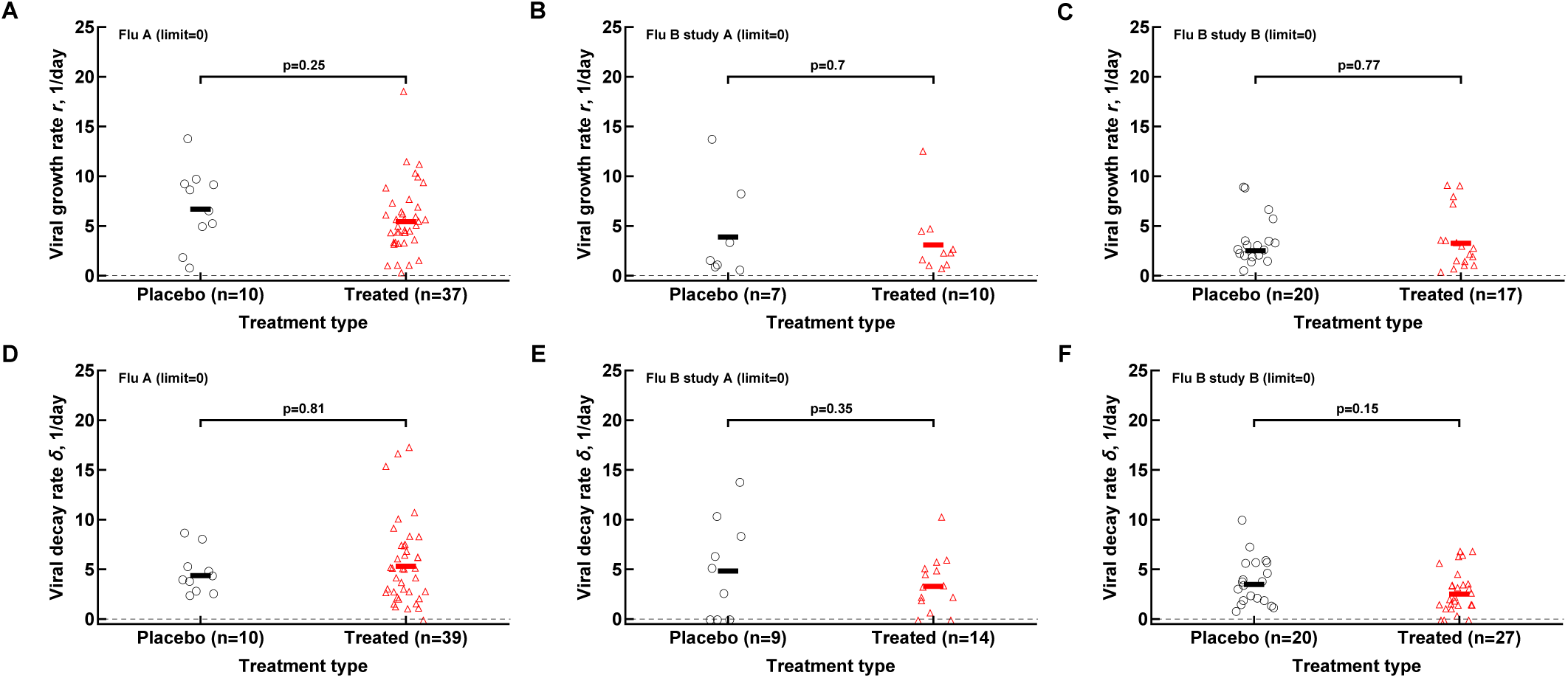
Oseltamivir treatment does not influence median viral growth and decline rates in individual volunteers. For the data from 3 clinical studies for every volunteer we calculated the rate at which viral shedding increased (A-C) or declined (D-F) with time for Flu A (A&D), Flu B study A (B&E), and Flu B study B (C&F) data. Only volunteers with the data above limit of detection (LOD) and at least two data points for virus increase to the peak or virus decline after the peak were included. Horizontal lines denote median values. The number of volunteers, *n*, analyzed in each of the trial and tests is shown on the *x*-axis of each graph. Comparisons between groups done using Mann-Whitney test and *p* values from the test are shown on individual panels.

In calculating virus growth and decay rates for individual volunteers we only used values that were above the LOD; therefore, this approach is likely to capture the average rates of virus shedding change. We wondered if drug treatment may impact the very early rate of virus shedding increase (or the very late virus shedding decline). Therefore, we also calculated two additional rates -the rate at which virus shedding increases early during the infection (by using the data that included one measurement at the LOD prior to detectable virus but before the viral peak) and the late rate of virus decline (by using the data after the peak shedding until the first measurement at LOD; see Materials and Methods for more detail). Interestingly, there was no evidence that the rate at which virus shedding increased early was changing over time either in placebo- or drug-treated volunteers (Figure S11). In contrast, the rate at which virus shedding was declining was slowing down with time since infection for some data (Figure S12).

Both flu A study and flu B study A tested the impact of different drug dosing on viral control (see Materials and Methods). Additional analysis showed that interestingly the oseltamivir’s dose had no measurable impact on the basic parameters for virus shedding kinetics which further justifies considering drug-treated volunteers as a single group (Figures S13 and S14).

### Matching median virus shedding curves with published studies

Original reports of the analyzed clinical trials included median viral shedding curves for the influenza A trial [12] and the influenza B study B trial [13]. We therefore investigated whether median viral shedding curves from our digitized data would accurately match published median viral curves. In our analyses we found that the assumed LOD influenced the resulting median shedding curve, and interestingly, by assuming LOD = 0 we found a nearly perfect match between median viral shedding in our data and published values for flu A clinical trial (Figure 5A&C). For the full match the median shedding data found in our analysis had to be shifted by 12h, though. In contrast, we could not fully match median viral shedding curves for flu B study B clinical trial for several values of the LOD (results not shown). Furthermore, even to provide a reasonable match with LOD = 0 the median shedding curve in our analysis had to be shifted by 24 hours (Figure 5B&D). We could not identify the reasons for this discrepancy. This result further suggests the need to publish raw data from such clinical trials so that some of the results can be independently verified.

**Figure 5:**
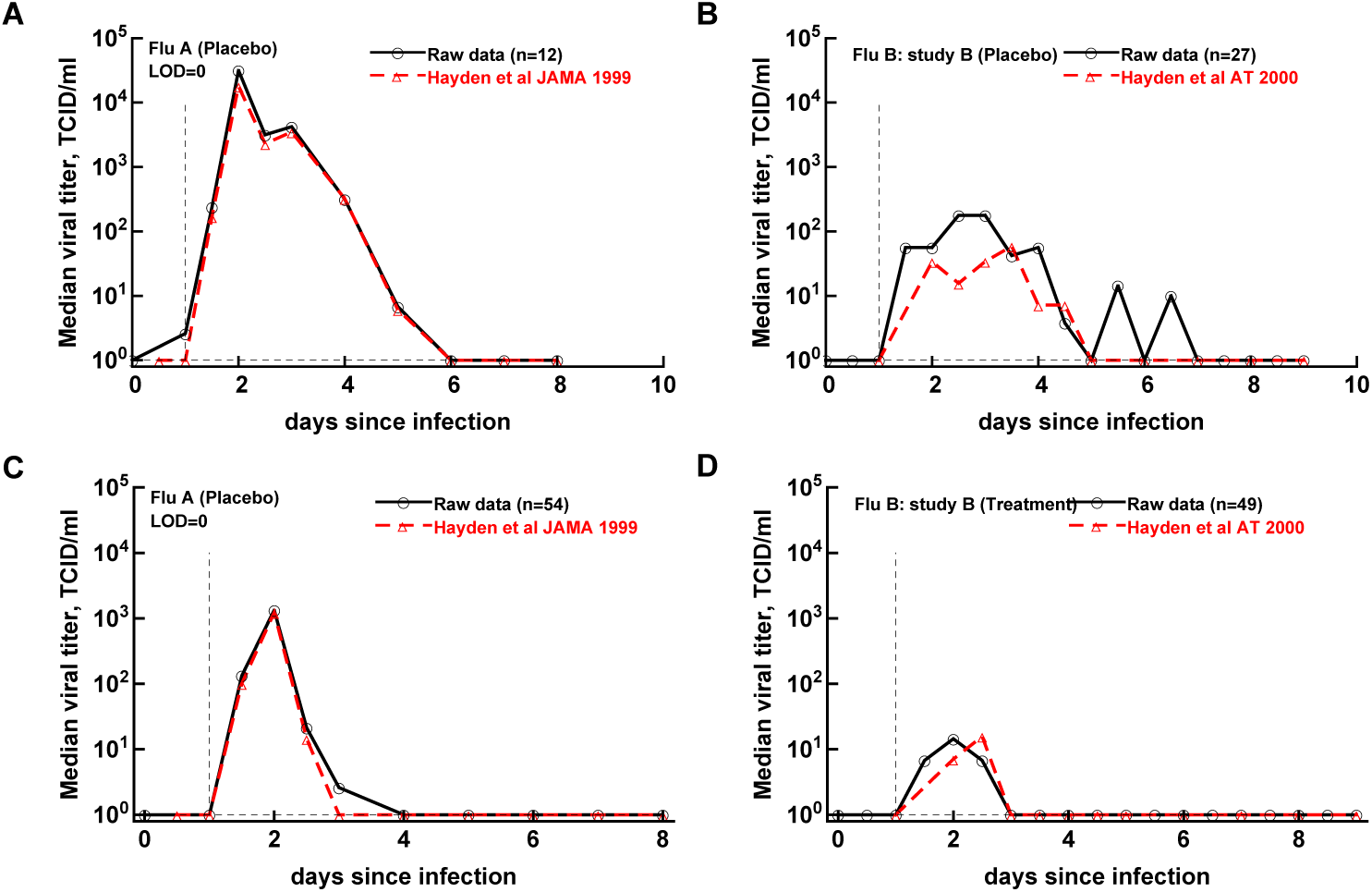
Previously published median virus shedding data could be reproduced for one but not another clinical trial. We compared median shedding data published previously for clinical trial with flu A [12] or flu B study B [13] with median shedding calculated from the raw trial data. The best match was found by assuming LOD = 0. For every panel we also list the number of volunteers used to calculate the median titers. The best match was obtained by shifting the published median shedding curves by 12 h (A&C) or by 24 hours (B&D).

### Oseltamivir treatment impacts the overall dynamics of start and end of shedding

Viral shedding dynamics in individual volunteers is relatively asynchronous – the time when individuals start or stop viral shedding (i.e., with viral shedding being above the LOD) varied. We therefore investigated if the rate at which volunteers started or stopped shedding was dependent on the virus type and, more importantly, on the oseltamivir treatment. We developed a novel mathematical model predicting start (or end) of shedding in the cohort of patients, based on a mixture of two gamma distributions, and estimated the model parameters by fitting a series of nested models to the data. Specifically, placebo- or drug-treated volunteers were followed as a cohort and the fraction of volunteers that started virus shedding by time *t* since infection (Figure 6A,C,E) or stopped shedding by time *t* (Figure 6B,D,F) was calculated. The models were fitted using a likelihood method and the best fit model was determined using likelihood ratio test (see Materials and methods for more detail and Figure 6).

**Figure 6:**
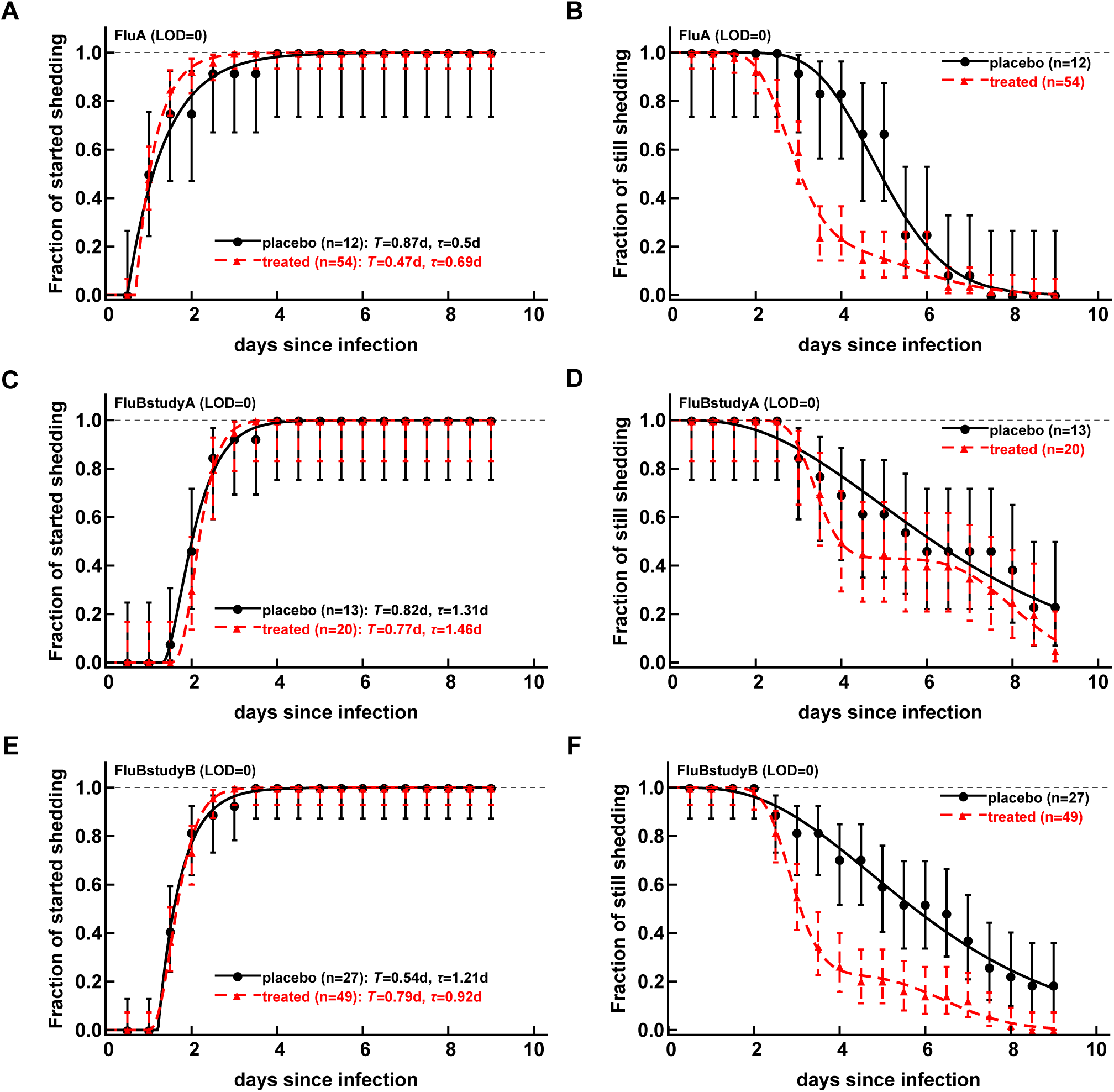
Oseltamivir treatment influences start and end of influenza virus shedding. We calculated the cumulative fraction of volunteers that start (A, C, E) or stop (B, D, F) shedding following infection with influenza A (A-B) or influenza B (C-E) viruses. Data are shown by markers for placebo or treated volunteers and in parentheses we show the numbers of volunteers in each cohort. We fitted a general mathematical model (given in eqn. (2) or eqn. (3)) to these data and the predictions of the best fit models are shown by lines. For every fit the minimal number of fitted parameters was selected using likelihood ratio test. Best fit parameters are shown in Table 3. In panels, *T* = *k/s*_*i*_ is the average shedding time and *τ* is the delay in shedding (see eqn. (2) for parameter definition). There was a moderate difference in the kinetics of start of shedding in Flu A study between placebo- and drug-treated volunteers (*p* = 0.046), but no difference was observed in start of virus shedding two studies with flu B. In FluA study and FluB study B the kinetics of stop of shedding was different between placebo and treated volunteers as judged by the likelihood ratio test (*p* < 0.001 for both comparisons); for FluB study A and study B there was not difference in kinetics of end of viral shedding (*p >* 0.2 for two comparisons). Error bars were estimated for binomial proportions using Jefferey’s intervals [31].

Several interesting results emerged. The kinetics of start of viral shedding was similar for both influenza virus types with half of individuals starting shedding within 1-2 days since infection. The best fit model describing shedding kinetics varied with the study and was dependent on the treatment type; interestingly, oseltamivir-treated volunteers infected with influenza A virus started shedding the virus with a different kinetics as compared to controls (LRT: *p* = 0.046, Figure 6A and Table 3). However, in other two studies start of shedding kinetics was similar between placebo- and drug-treated volunteers (Figure 6 and Table 3).

**Table 3:** Parameters providing the best fit of the model (given by eqn. (2) or eqn. (3)) to the data on start or end of shedding by volunteers in three clinical trials. Data and model fits are show in Figure 6. To generate the survival curves we used LOD = 0. The models were fitted to the data from 3 clinical trials (Flu A, Flu B study A, and Flu B study B), for placebo or oseltamivir-treated volunteers, and for start or end of shedding. Each dataset is thus named to include these details. When values are given as integers (e.g., *s*_2_ = 0, *f* = 1, *k* = 1, or *τ* = 0), these parameters were fixed to the noted values in model fits to data. Allowing these parameters to be fit did not improve the quality of the model fit of the data based on likelihood ratio test (results not shown). Confidence intervals in estimated parameters (shown in parentheses) were estimated by resampling data for individual volunteers 1,000 times with replacement.

| Study | Treatment | Shedding | $s_1$, 1/day | $s_2$, 1/day | $f$ | $k$ | $\tau$, day |
| --- | --- | --- | --- | --- | --- | --- | --- |
| Flu A | placebo | start | 1.16 (0.78-34.64) | 0 | 1 | 1 | 0.5 (0.5-0.96) |
| Flu A | treated | start | 2.15 (1.62-3.69) | 0 | 1 | 1 | 0.69 (0.52-0.83) |
| Flu A | placebo | stop | 3.35 (1.68-15.27) | 0 | 1 | 16.79 (8.61-84.87) | 0 |
| Flu A | treated | stop | 3.27 (0.94-4.08) | 6.72 (2.47-8.55) | 0.2 (0.07-0.61) | 19.36 (7.66-23.64) | 0 |
| Flu B study A | placebo | start | 2.48 (0.98-26.85) | 0 | 1 | 2.03 (0.2-48.42) | 1.31 (0-2.0) |
| Flu B study A | treated | start | 5.68 (1.85-46.24) | 0 | 1 | 4.39 (0.44-74.43) | 1.46 (0-1.99) |
| Flu B study A | placebo | stop | 0.54 (0.2-0.96) | 0 | 1 | 3.63 (1.93-5.72) | 0 |
| Flu B study A | treated | stop | 6.48 (1.87-13.67) | 15.44 (5.29-36.51) | 0.43 (0.2-0.64) | 52.75 (18.61-124.6) | 0 |
| Flu B study B | placebo | start | 1.87 (1.35-3.75) | 0 | 1 | 1 | 1.21 (1.0-1.39) |
| Flu B study B | treated | start | 4.91 (3.30-14.37) | 0 | 1 | 3.9 (2.15-32.62) | 0.92 (0.0-1.0) |
| Flu B study B | placebo | stop | 0.66 (0.30-1.02) | 0 | 1 | 4.12 (2.18-6.47) | 0 |
| Flu B study B | treated | stop | 4.72 (3.07-8.10) | 10.73 (7.11-21.54) | 0.23 (0.12-0.36) | 31.3 (20.32-57.77) | 0 |

The kinetics at which volunteers stopped shedding was even more intriguing. Only for flu B study A we found no differences in the kinetics of viral shedding end between placebo- and oseltamivir-treated volunteers (Figure 6D); however, the model fits predicted that in about 43% of drug-treated individuals the loss of virus shedding proceeded similarly to that of placebo-treated volunteers (given by parameter *f* in Table 3). Indeed, average time to stop shedding was 3.63*/*0.54 = 6.7 days and 52.75*/*6.48 = 8.1 days for placebo- and drug-treated volunteers, respectively, suggesting that the drug may have failed in preventing virus replication in many individuals in this trial. There were statistically significant differences in the kinetics of loss of viral shedding between placebo- and drug-treated volunteers in flu A study and flu B study B (Figure 6B&F and Table 3). In these trials, about 20% of drug-treated volunteers stopped shedding the virus with similar timing to that of placebo-treated individuals (Table 3). Thus, our novel analysis identified variable impact of the oseltamivir treatment on the kinetics at which virus shedding starts or ends in human volunteers.

## Discussion

One of the great advantages of testing efficacy of vaccines or drugs against influenza viruses is the ability to perform controlled human challenge studies in which volunteers (placebo and drug-treated or vaccinated) are exposed to well defined dose and type of the virus [32, 33]. However, results of such important trials in general have been presented in succinct manner; e.g., typically median virus shedding curves are presented and public access to data on influenza virus dynamics in individual volunteers is not provided [12, 13, 18]). In our personal experience researchers that do have access to such raw data have not been willing to share them. In this study we took advantage of now publicly available reports from three clinical studies testing efficacy of oseltamivir given 24 hours after exposure volunteers to influenza A or B viruses. We have converted scanned pdf pages into spreadsheet format data that can be analyzed further and make it a resource for the community. We have also performed several basic analyses of these data.

We found that some of the previously published results could be reproduced, i.e., oseltamivir treated did reduce the duration of viral shedding and the overall viral shedding in 2 out of 3 trials (Figure 3); however, a closer inspection of the data revealed that many treated volunteers continued shedding the virus suggesting treatment failure. Indeed, by looking at the kinetics at which volunteers stop shedding we found that between 20-40% of treated volunteers continued shedding similarly as placebo-treated controls further suggesting treatment failure (Figure 6). Our mathematical model-driven analysis also suggested that treatment with oseltamivir may in fact speed up start of shedding, most noticeable in flu A clinical trial (Figure 6A). While mechanisms of this process are unclear – we could not find information on the volunteers that shed the virus early – further studies may need to investigate this possibility further.

We could only partially reproduce previously published median shedding curves, specifically for flu A trial but only after shifting the published median shedding curves to allow for a nearly perfect match (Figure 5A&C). Why it was not possible for flu B study B remains unclear. It is also interesting to note that while analysis of the median shedding curves in flu A study suggested a faster viral clearance in oseltamivir-treated volunteers [22], virus clearance rates were similar when evaluated for individual volunteers (Figure 4). Given that oseltamivir treatment also did not impact the early virus growth rate this may suggest that virus shedding is controlled mainly by innate mechanisms (e.g., target cell limitation, [19, 22]). Further studies should investigate in more detail whether alternative models for virus control are consistent with shedding data for individual volunteers infected with influenza A or B viruses [22, 34].

Our work has some limitations. Because original virus shedding data were in PDF format we cannot exclude the possibility that there have been errors in our translation from the given days/times since infection to digital format. We attempted to minimize such errors by having another investigator to check correctness of the digitization. We could not determine why some volunteers responded well to the drug and improved clearance of the virus while others did not. It was not possible to find clear answers in the clinical trial notebooks. Our analysis of virus growth and decay rates depended strongly on the measured values (above LOD), and more frequent measurements of viral shedding are likely to provide a more accurate estimates of these rates.

Our work generates several recommendations that should be implemented for clinical trial data on human challenge studies with influenza viruses. We believe that original shedding data should be shared following publication of the work. The data can be anonymized and redacted if needed, but the critical numeric data must be provided in the proper digital format (e.g., spreadsheets or similar). For example, recent clinical trial data for baloxavir have not been made available [18]. Authors that provide re-analysis of the data from clinical trials should also attempt to provide public access to such data. Data sharing is likely to improve science reproducibility which is likely to ultimately benefit influenza virus-infected patients.

## Data Availability

Data will be available following publication of the paper in peer reviewed literature as supplement.

## Author contributions

KLH digitized the data from original publications into spreadsheet format. KLH and VVG have analyzed the data and wrote the manuscript.

## Data sources

Data on virus shedding from the three clinical studies are available as supplement to this paper.

## Code sources

Mathematica codes used for data analysis are available as supplement to this paper.

## Acknowledgments

We would like to thank our research community for pressuring Roche to release primary research data from oseltamivir clinical trials. We also thank several students in the GanusovLab for helping with some primary analyses and checks of the digitized data. This work was in part supported by the NIH grant (R01 GM118553) to VVG.

## Abbreviations

LOD: limit of detection
DOI: days of infection (also denoted as *T*)
bid: twice daily
odd: once daily
AUC: area under the curve
LRT: likelihood ratio test.

## Supplemental information

**Figure S1:**
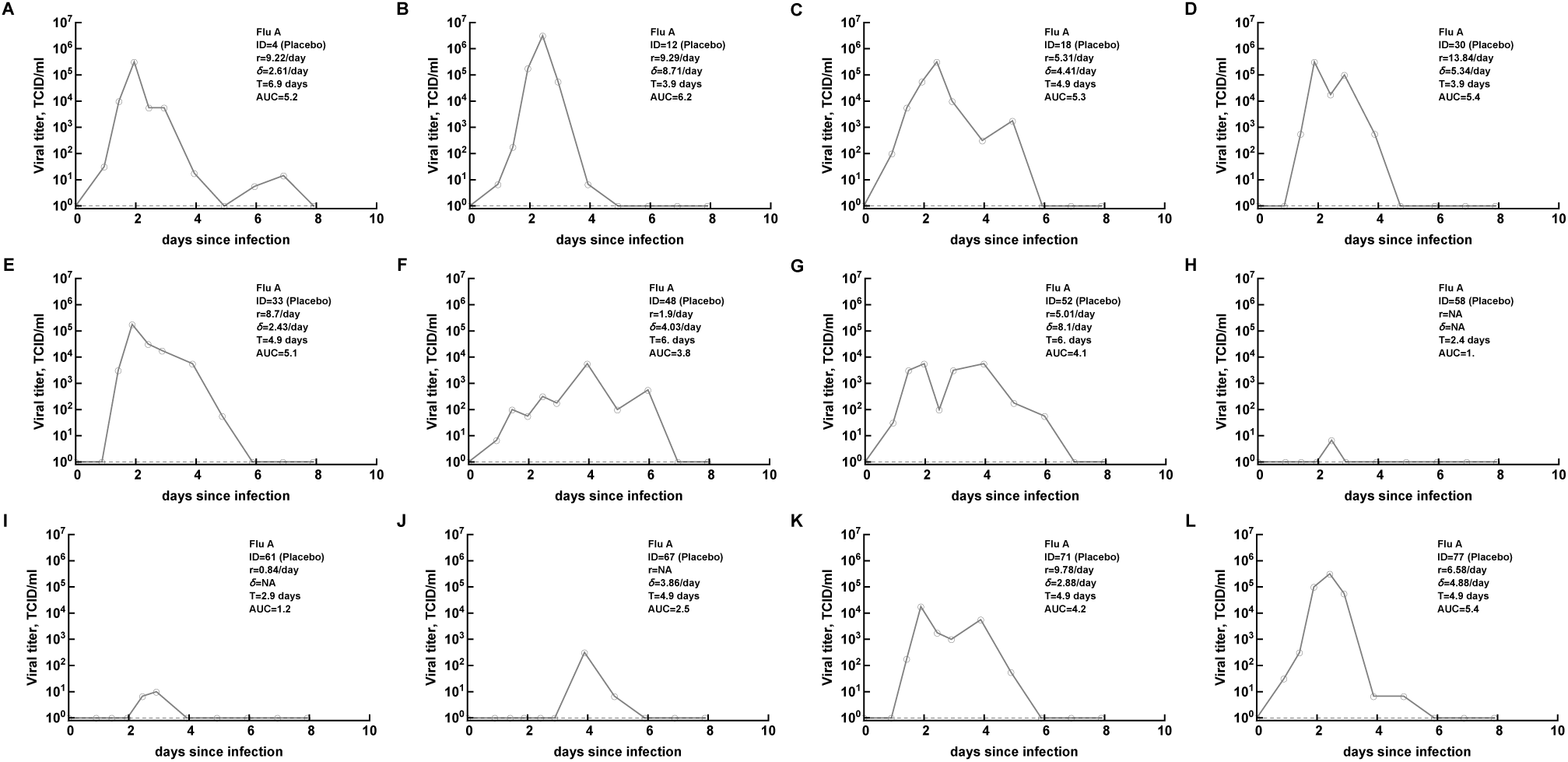
Viral shedding titers for individual volunteers from Flu A study [12]. These are data for placebo-treated volunteers including the volunteer ID, duration of infection, viral growth and viral decline rates, duration of infection, and the total viral sheeting (AUC). Volunteers excluded from the analysis as uninfected have the following IDs: 7, 21, 38, 41.

**Figure S2:**
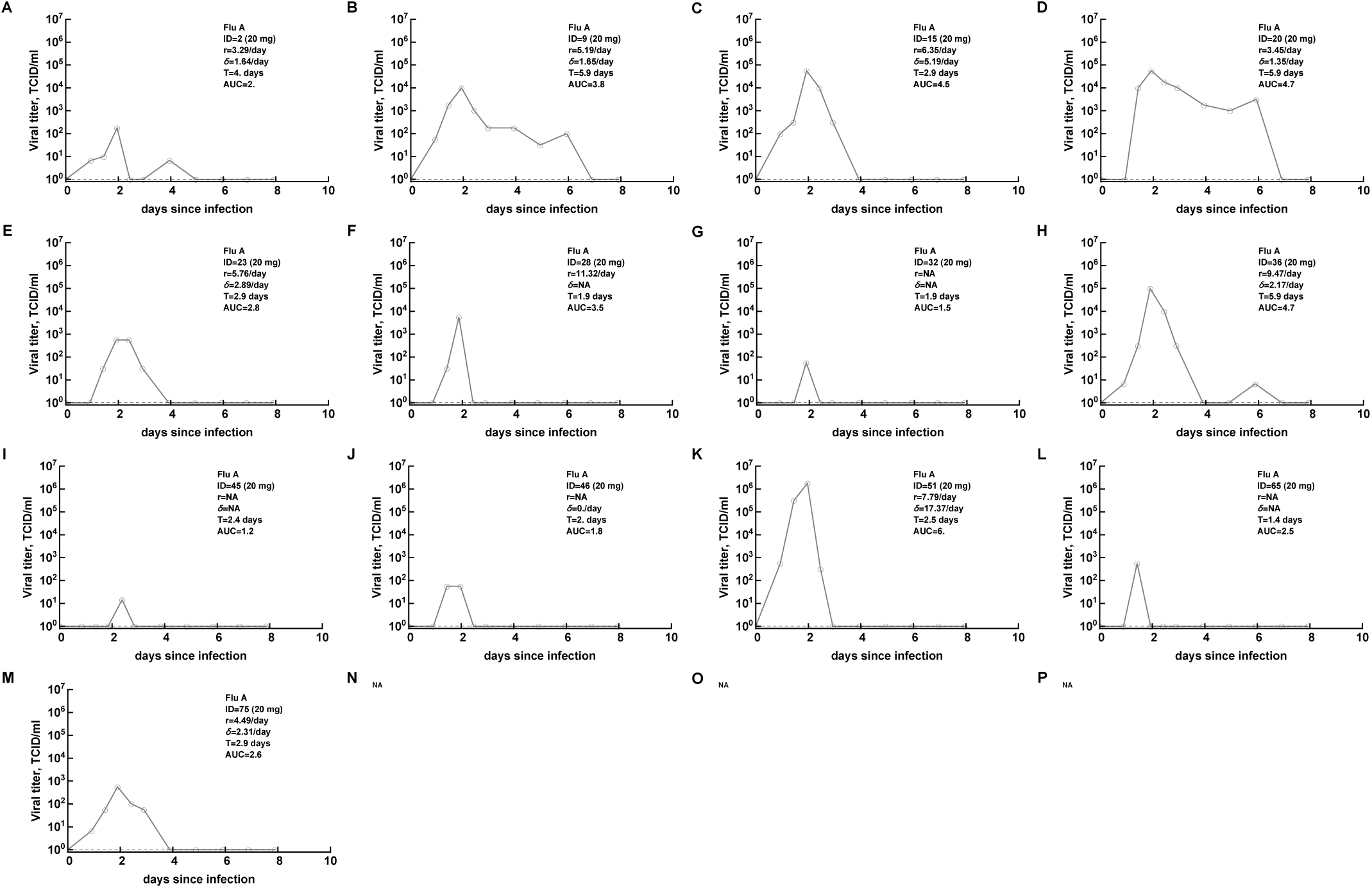
Viral shedding titers for individual volunteers from Flu A study [12] treated with 20mg of oseltamivir. Volunteers excluded from the analysis as uninfected have the following IDs: 56, 68, 79. See Figure S1 for more detail.

**Figure S3:**
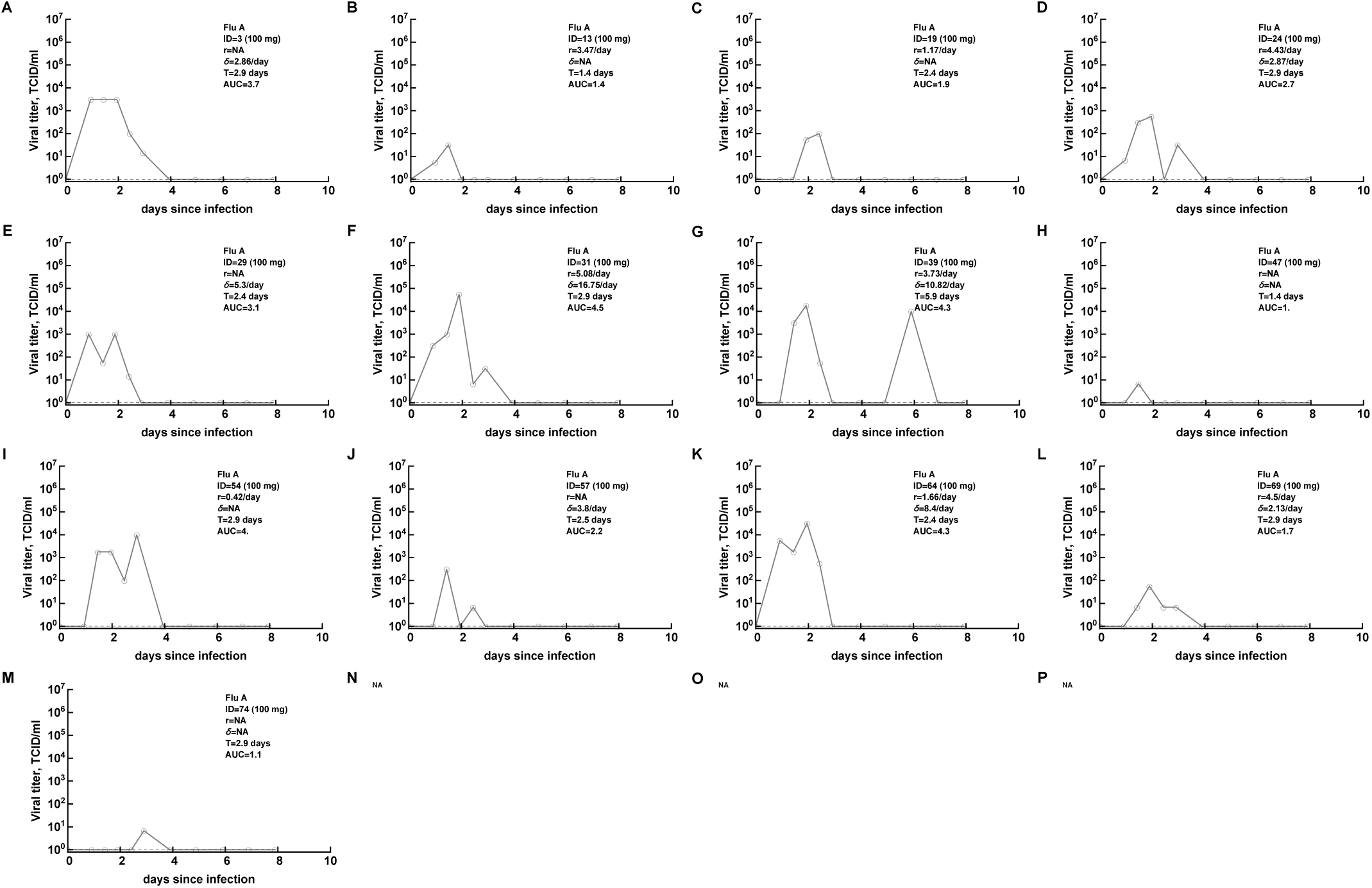
Viral shedding titers for individual volunteers from Flu A study [12] treated with 100mg of oseltamivir. Volunteers excluded from the analysis as uninfected have the following IDs: 10, 42, 78. See Figure S1 for more detail.

**Figure S4:**
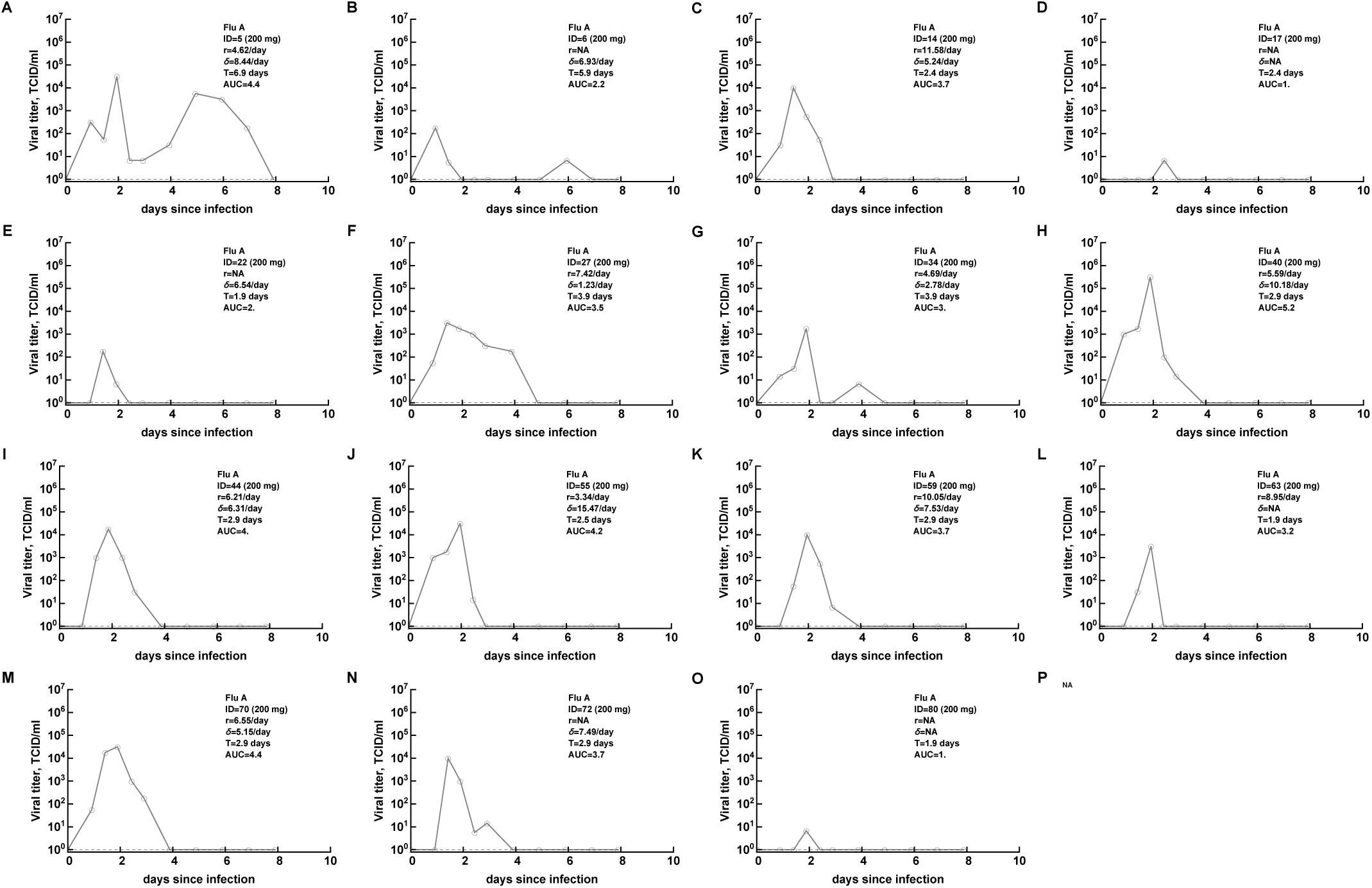
Viral shedding titers for individual volunteers from Flu A study [12] treated with 200mg of oseltamivir twice daily. Volunteers excluded from the analysis as uninfected have the following IDs: 50. See Figure S1 for more detail.

**Figure S5:**
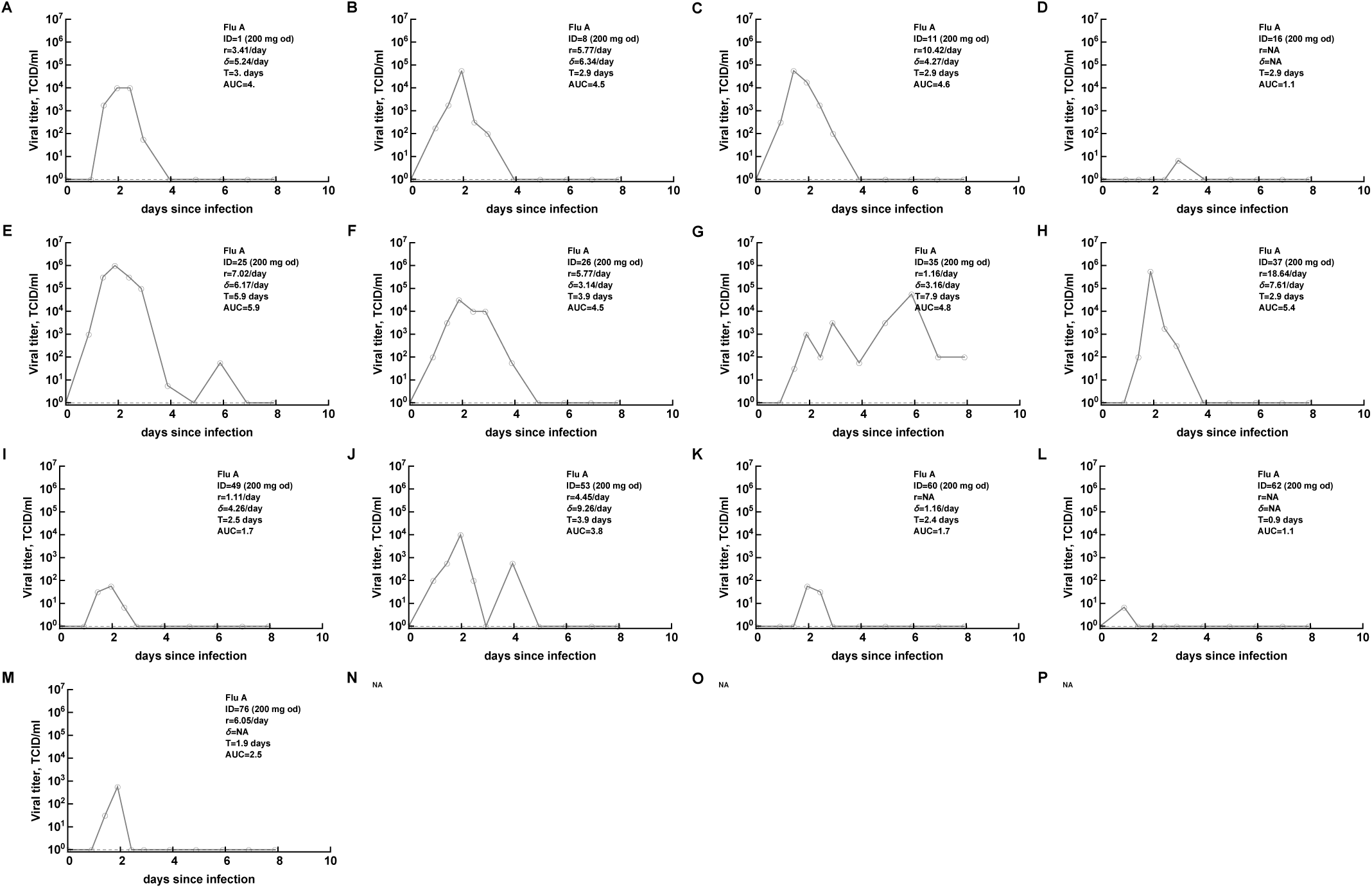
Viral shedding titers for individual volunteers from Flu A study [12] treated with 200mg of oseltamivir once daily. Volunteers excluded from the analysis as uninfected have the following IDs: 43, 66,73. See Figure S1 for more detail.

**Figure S6:**
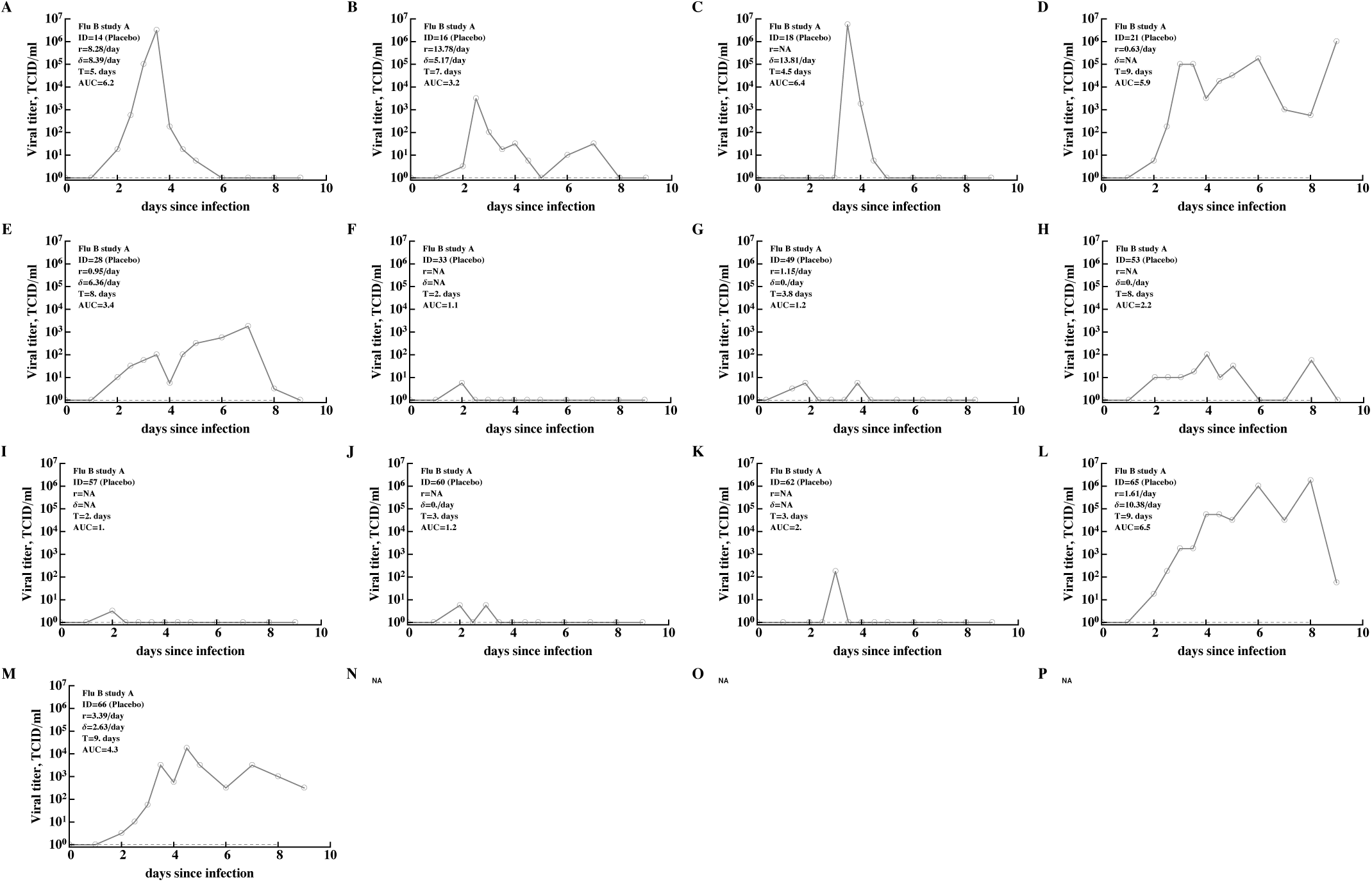
Viral shedding titers for individual volunteers from Flu B study A [13]. These are data for placebo-treated volunteers including the volunteer ID, duration of infection, viral growth and viral decline rates, duration of infection, and the total viral sheeting (AUC). Volunteers excluded from the analysis as uninfected have the following IDs: 25, 31, 35, 36, 41, 44, 47.

**Figure S7:**
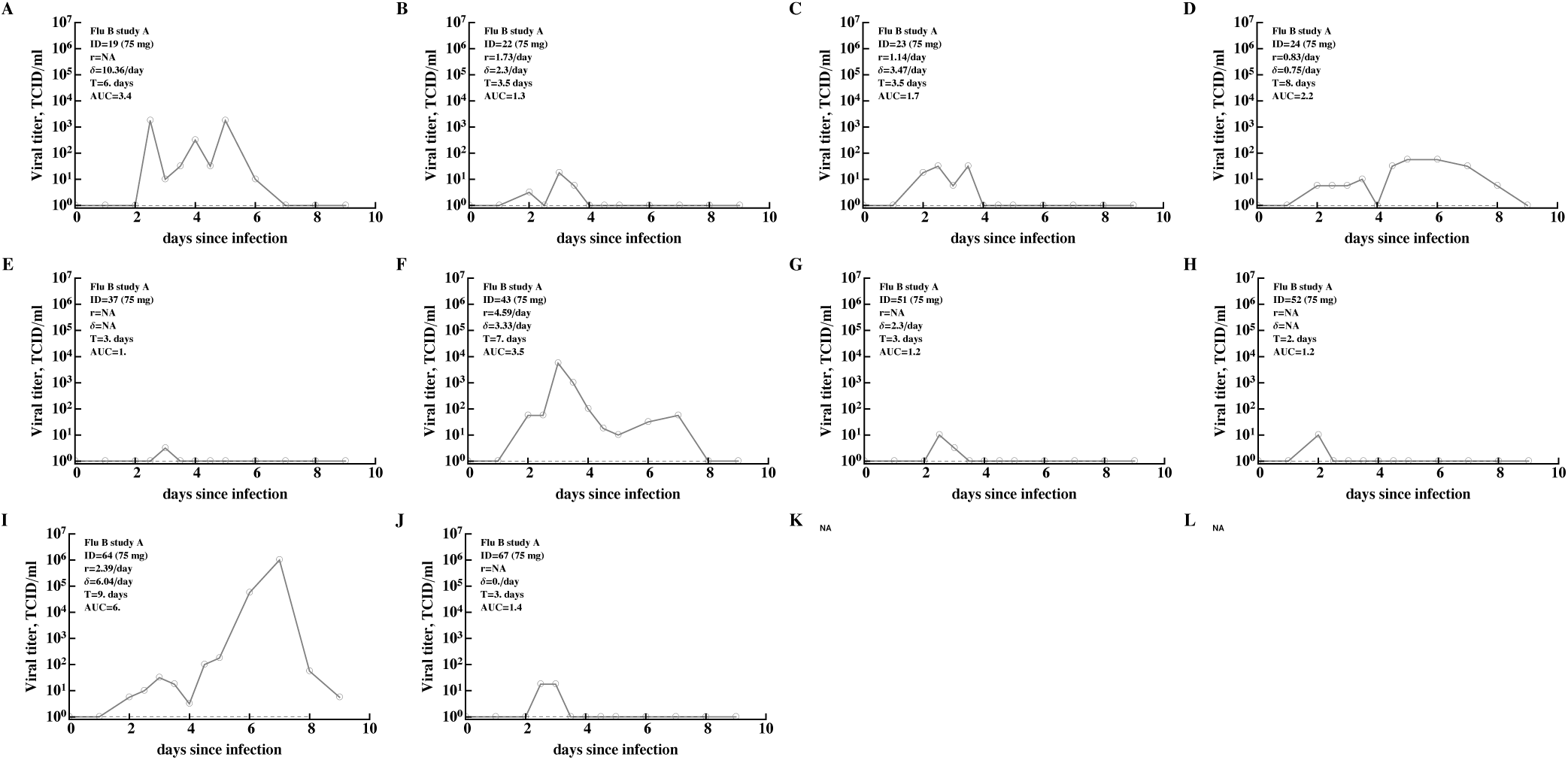
Viral shedding titers for individual volunteers from Flu B study A [13] treated with 75mg of oseltamivir. Volunteers excluded from the analysis as uninfected have the following IDs: 12, 13, 30, 32, 39, 45, 55, 58, 59, 70. See Figure S6 for more detail.

**Figure S8:**
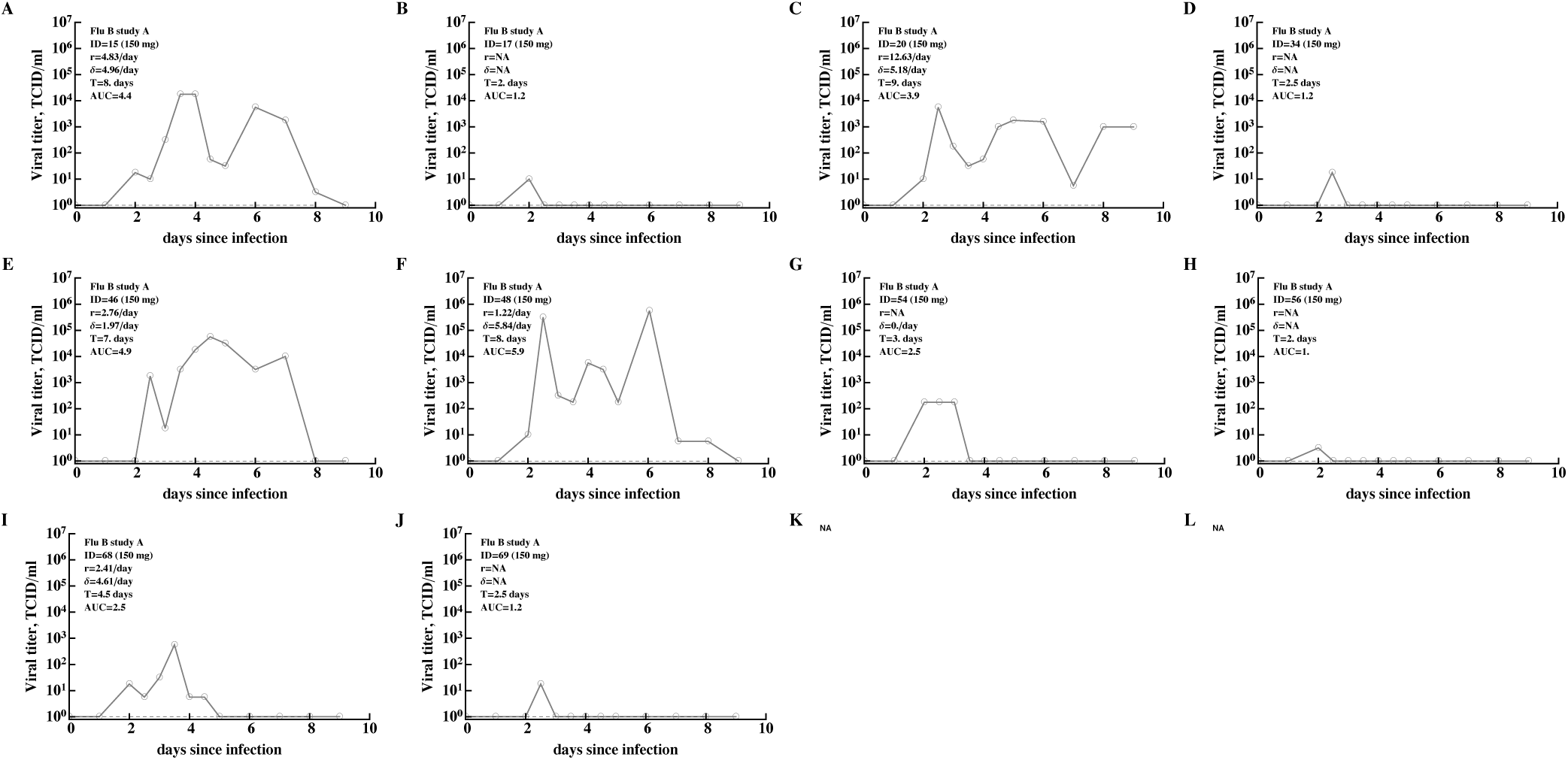
Viral shedding titers for individual volunteers from Flu B study A [13] treated with 150mg of oseltamivir. Volunteers excluded from the analysis as uninfected have the following IDs: 11, 26, 27, 29, 38, 40, 42, 50, 61, 63. See Figure S6 for more detail.

**Figure S9:**
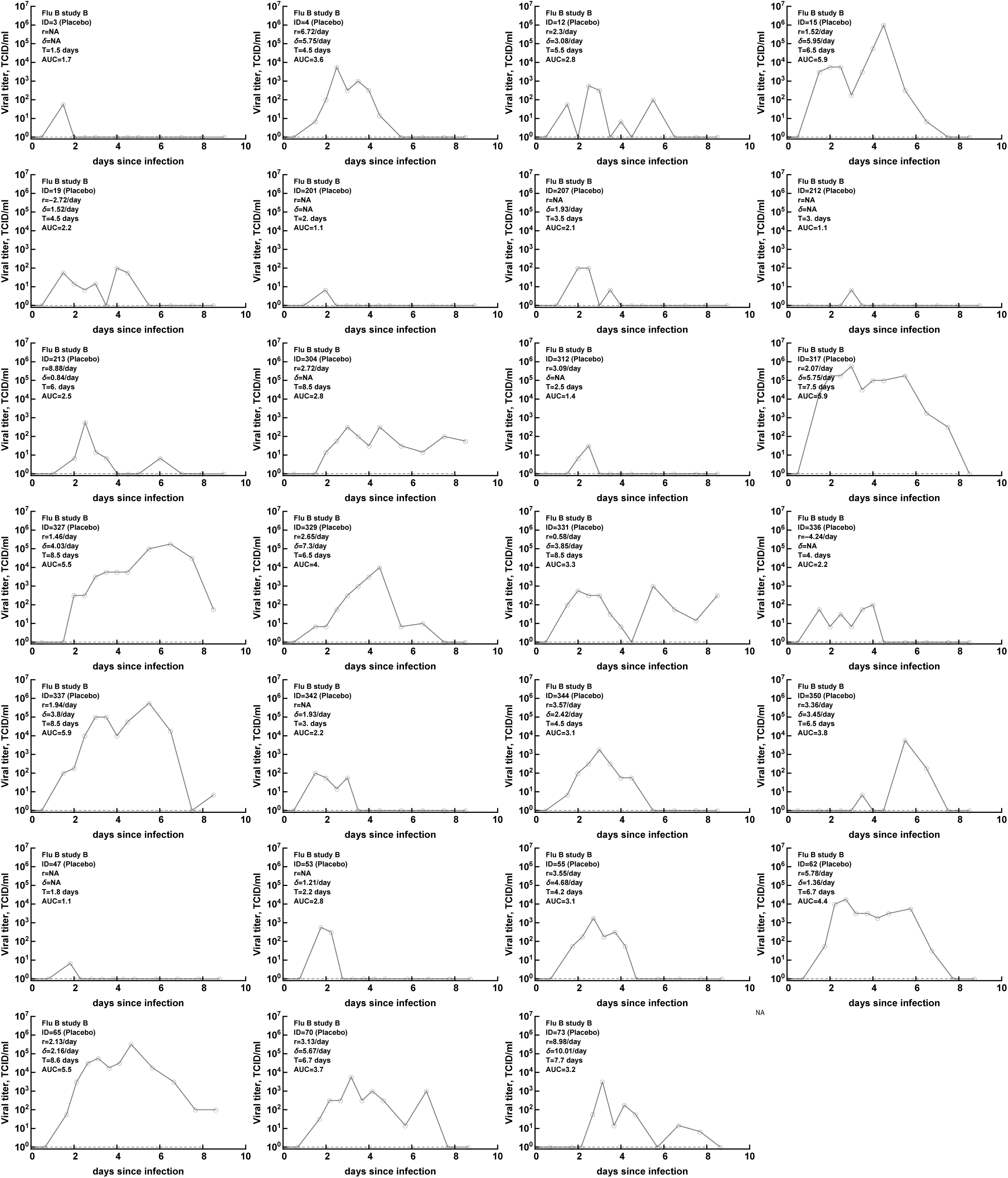
Viral shedding titers for individual volunteers from Flu B study B [13]. These are data for placebo-treated volunteers including the volunteer ID, duration of infection, viral growth and viral decline rates, duration of infection, and the total viral sheeting (AUC). Volunteers excluded from the analysis as uninfected have the following IDs: 8, 17, 204, 303, 308, 314, 319, 324, 346, 49, 58, 67.

**Figure S10:**
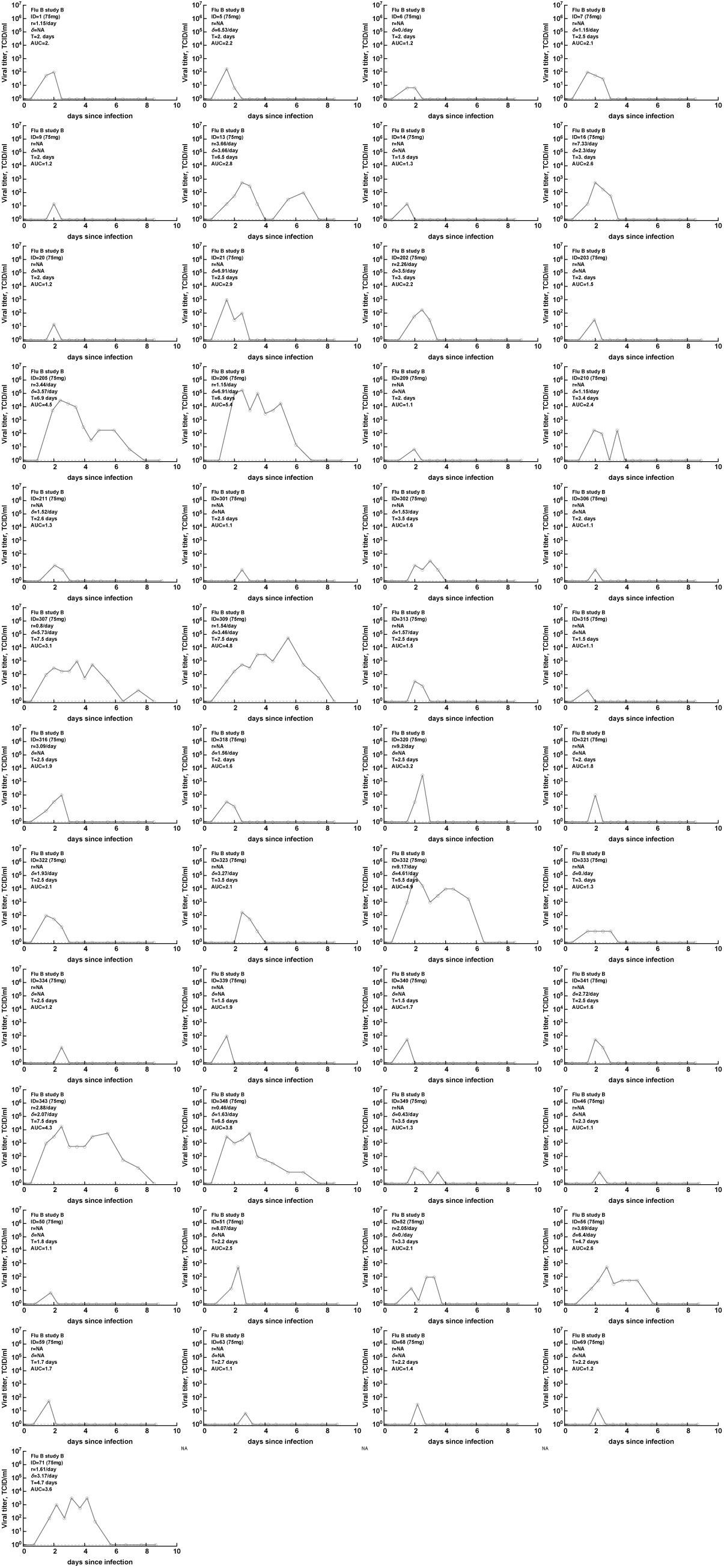
Viral shedding titers for individual volunteers from Flu B study A [13] treated with 75mg of oseltamivir. Volunteers excluded from the analysis as uninfected have the following IDs: 2, 10, 11, 18, 208, 214, 215, 305, 310, 311, 325, 326, 328, 330, 335, 338, 345, 347, 351, 48, 54, 57, 60, 61, 64, 66, 72, 74, 75. See Figure S9 for more detail.

**Figure S11:**
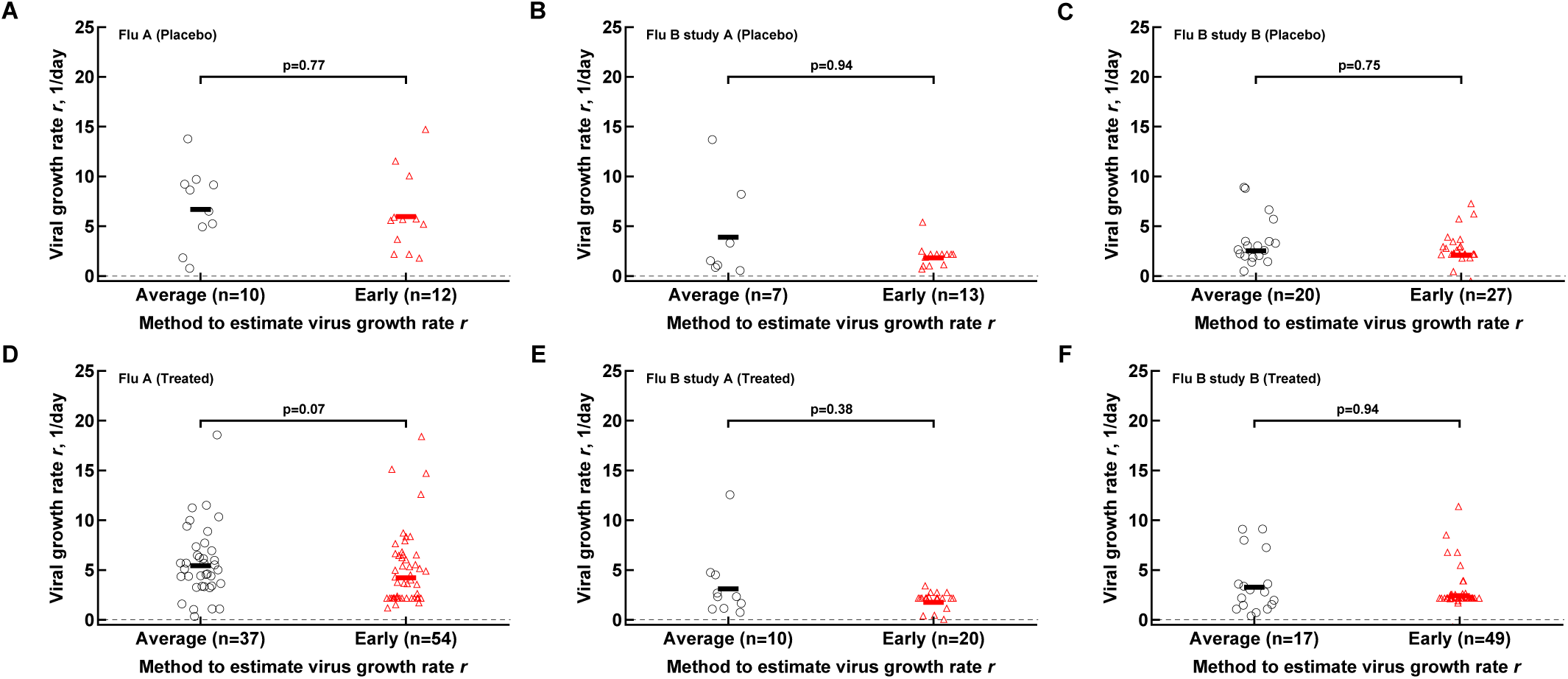
Oseltamivir treatment does not result in acceleration of early viral replication rates. We calculated the early virus replication rate by either only including viral shedding data prior to peak that are above the limit of detection (“Average” category) or by including all data points prior to peak including data at the limit of detection (“Early” category). Panels A&D show the Influenza A analyses. Panels B&E show the Influenza B study A analyses. Panels C&F show the Influenza B study B analyses. The small black line in each of the columns of data show the median value for that analysis. The number of volunteers, *n*, analyzed in each of the trial and tests is shown on the x-axis of each graph. *P*-values were calculated using the Mann-Whitney test.

**Figure S12:**
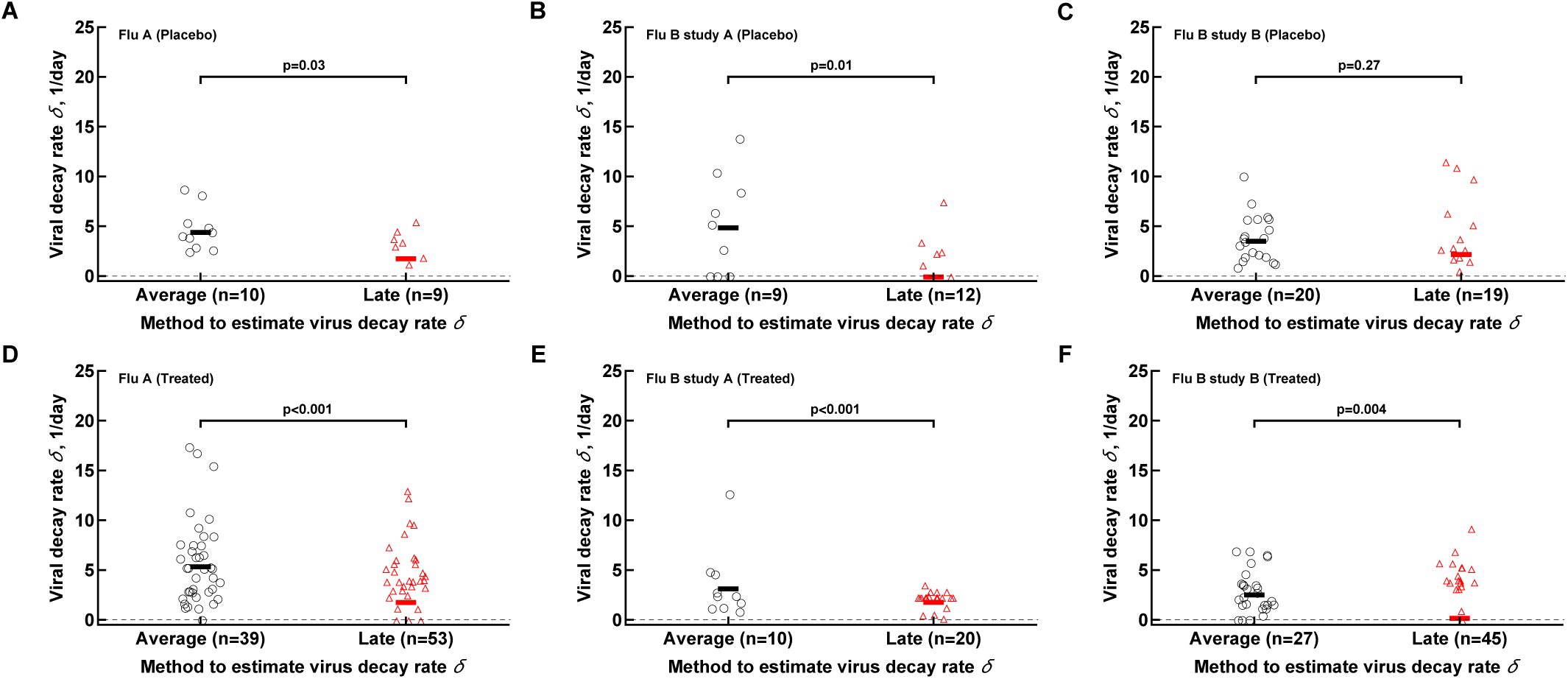
Oseltamivir treatment results in a slower viral clearance over time. We calculated the late virus decline rate by either only including viral shedding data after the peak that are above the limit of detection (“Average” category) or by including all data points after the peak including one first data point at the limit of detection (“Late” category). Panels A&D show the Influenza A analyses. Panels B&E show the Influenza B study A analyses. Panels C&F show the Influenza B study B analyses. The small black line in each of the columns of data show the median value for that analysis. The number of volunteers, *n*, analyzed in each of the trial and tests is shown on the x-axis of each graph. P-values were calculated using the Mann-Whitney test.

**Figure S13:**
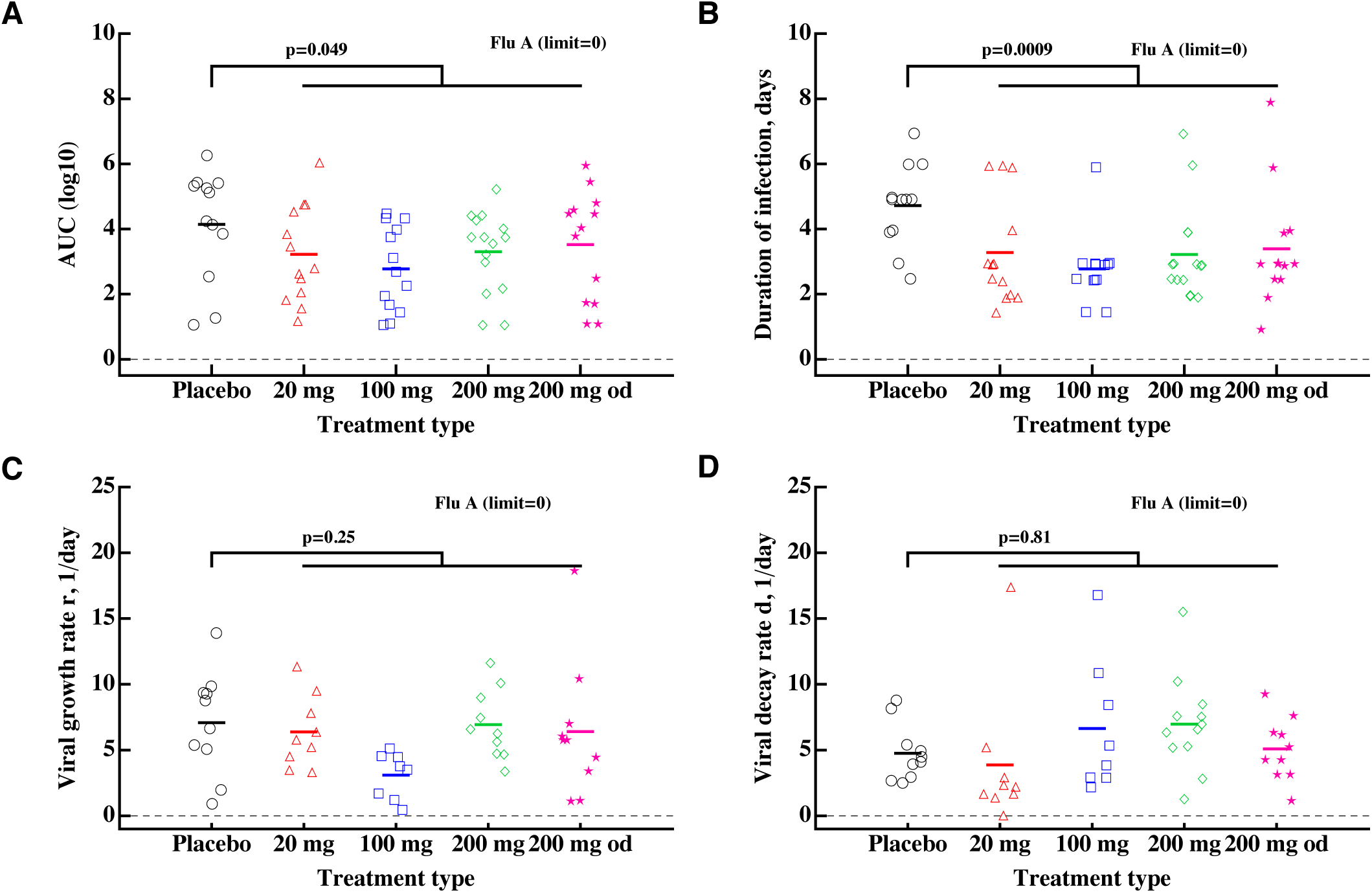
Impact of oseltamivir treatment type (dose) on the total viral shedding (AUC, A), duration of infection (B), viral growth (C) and decline (D) rates for individuals in Flu A clinical trial [12]. The differences in estimated parameters between different treatments were not significant (judged by ANOVA). See Figures 3 and 4 for more detail.

**Figure S14:**
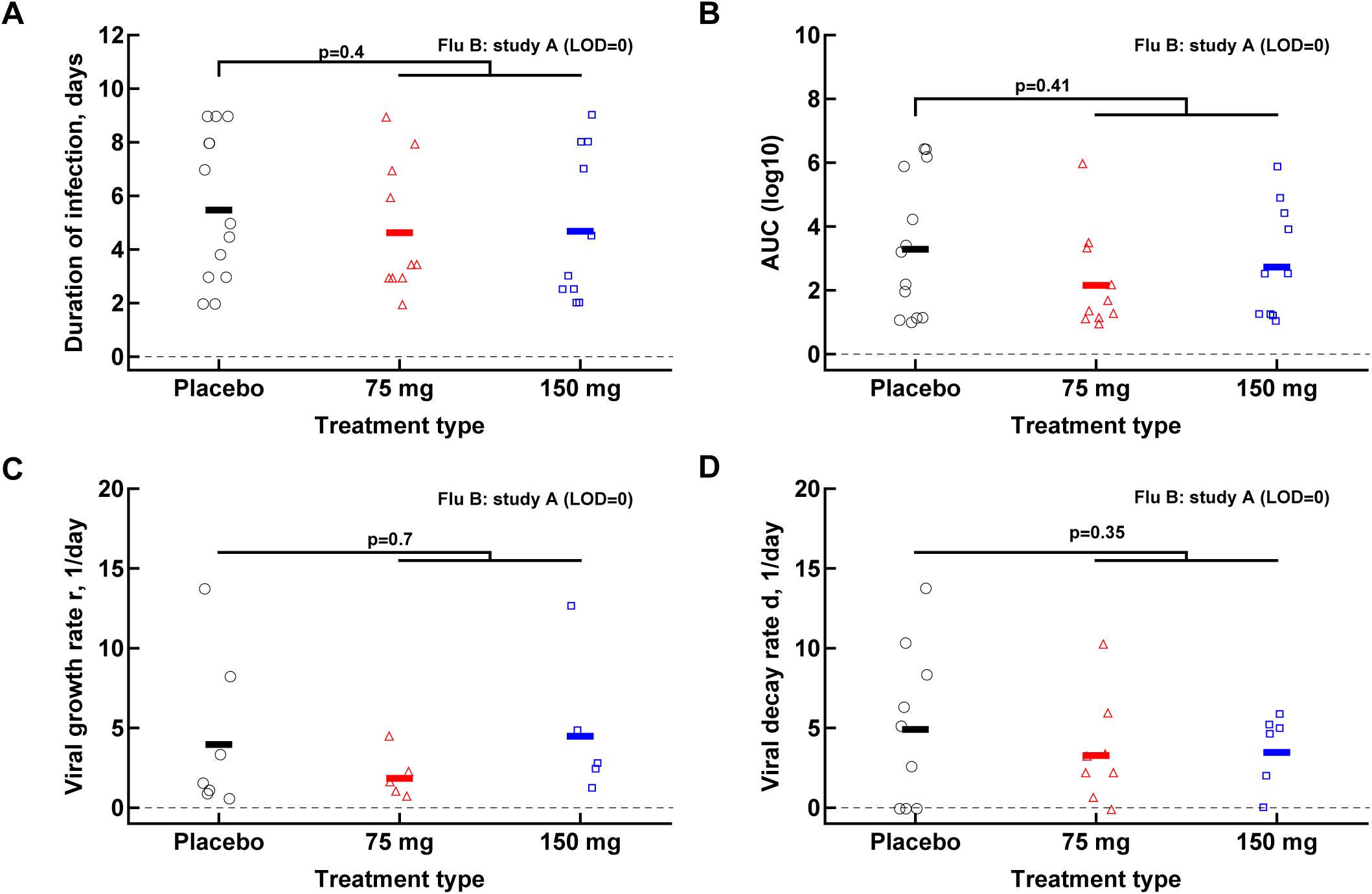
Impact of oseltamivir treatment type (dose) on the total viral shedding (AUC, A), duration of infection (B), viral growth (C) and decline (D) rates for individuals in Flu B study A clinical trial [13]. The differences in estimated parameters between different treatments were not significant (judged by ANOVA). See Figures 3 and 4 for more detail.

